# Diverging the anthracycline class of anti-cancer drugs for superior survival of acute myeloid leukemia patients

**DOI:** 10.1101/2023.11.23.23298950

**Authors:** Xiaohang Qiao, Sabina Y. van der Zanden, Xiaoyang Li, Minkang Tan, Yunxiang Zhang, Ji-Ying Song, Merle A. van Gelder, Feija L. Hamoen, Lennert Janssen, Charlotte L. Zuur, Baoxu Pang, Olaf van Tellingen, Junmin Li, Jacques Neefjes

**Affiliations:** Division of Tumor Biology and Immunology, The Netherlands Cancer Institute, Amsterdam, The Netherlands; Department of Head and Neck Oncology and Surgery, The Netherlands Cancer Institute, Amsterdam, The Netherlands; Department of Cell and Chemical Biology, ONCODE Institute, Leiden University Medical Center, Leiden, The Netherlands; Shanghai Institute of Hematology, State Key Laboratory of Medical Genomics, National Research Center for Translational Medicine, Ruijin Hospital affiliated to Shanghai Jiao Tong University School of Medicine, Shanghai, China; Division of Pharmacology, The Netherlands Cancer Institute, Amsterdam, The Netherlands; Division of Experimental Animal Pathology, The Netherlands Cancer Institute, Amsterdam, The Netherlands

**Keywords:** Anthracycline, Doxorubicin, Aclarubicin, Acute Myeloid Leukemia, Cardiotoxicity, Histone Eviction, Bio-distribution, Cross-resistance

## Abstract

The efficacy of anthracycline-based chemotherapeutics, which include doxorubicin and its structural relatives daunorubicin and idarubicin, remains almost unmatched in oncology, despite a side effect profile including cumulative dose-dependent cardiotoxicity, therapy-related malignancies and infertility. Detoxification of anthracyclines while preserving their anti-neoplastic effects is arguably a major unmet need in modern oncology, as cardiovascular complications that limit anti-cancer treatment are now a leading cause of morbidity and mortality among the 17 million cancer survivors in the U.S.. To address this, we examined different clinically relevant anthracycline drugs with respect to a series of features including mode of action (chromatin and DNA damage), bio-distribution, anti-tumor efficacy and cardiotoxicity in pre-clinical models and patients. We show that different anthracycline drugs have surprisingly individual efficacy and toxicity profiles. In particular, aclarubicin stands out in pre-clinical models and clinical trials as it potently kills cancer cells, does not induce therapy-related malignancies or cardiotoxicity, and can be safely administered even after a maximum cumulative dose of either ida- or doxorubicin has been reached. Retrospective analysis of aclarubicin used in second-line treatment of relapsed/refractory AML patients showed similar survival effects to its use in first line, leading to an almost 25% increase in 5-year overall survival. Considering individual anthracyclines as different drugs provides new treatment options that strongly improve survival of cancer patients while limiting the toxic side-effects.

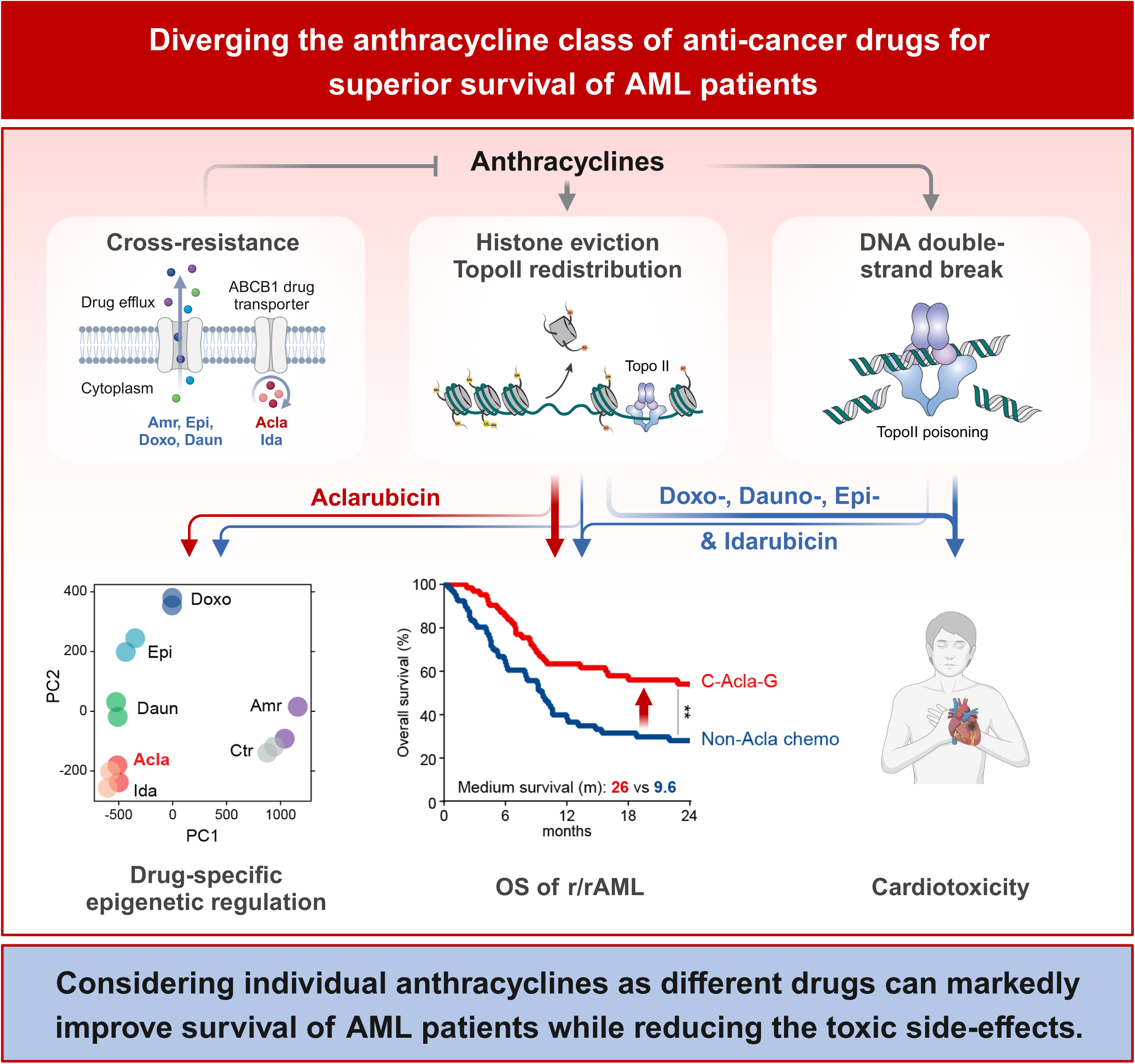

## Introduction

Most anthracyclines function as poisons of the enzyme topoisomerase II (TopoII) that trap it on chromatin and thus induce DNA double-strand breaks (DSBs) [1]. In addition, these drugs also evict histones from defined genomic areas, which results in delayed DNA damage repair and epigenetic and transcriptomic alterations, collectively termed chromatin damage [2–4]. Histone eviction appears to be a major cytotoxic activity of anthracyclines [5]. TopoII poisons, like etoposide, that only generate DSBs are considerably less effective in cancer treatment [2, 6], and also less toxic [5, 7]. On the contrary, aclarubicin only induces histone eviction and is highly effective in treating *de novo* acute myeloid leukemia (AML) [5]. Hence, DNA damage appears not to be a prerequisite for effective anti-cancer activities. Removing DNA damage from chromatin damage activity mitigates cardiotoxicity and therapy-related tumorigenesis of anthracyclines, without hampering their anti-cancer effect [5]. Clinical application of detoxified anthracyclines will confer major benefits in cancer therapy, expanding treatment options for patients with comorbidities including older adults [8] and reducing late effects, particularly for pediatric patients [9]. Detoxified anthracyclines would enable chronic treatment of cancer, while the current toxic anthracyclines only allow for limited treatment due to cumulative dose-dependent cardiotoxicity risk. Here, we systematically evaluated anthracycline drugs currently used in clinic against a series of key features to determine whether they differ in terms of anti-neoplastic potency, site of action and toxicity profiles. We identified aclarubicin, currently used only in Asia for AML patients with high comorbidity indices, as an anthracycline variant that lacks cardiotoxicity side effects and can be used after maximum dose of doxo- or ida in mice and men. Aclarubicin-based therapy increases the overall survival of relapsed/refractory AML patients by almost 25%. These data illustrate how aclarubicin can transform leukemia treatment for children, unfit adults and relapsed/refractory AML patients that currently have a poor prognosis, addressing a major unmet need in current oncology practice.

## Materials and methods

### Reagents

Doxorubicin and etoposide were purchased from Pharmachemie (The Netherlands), daunorubicin was obtained from Sanofi-Aventis and epirubicin was obtained from Accord Healthcare Limited (UK), idarubicin was obtained from Pfizer and Santa Cruz Biotechnology (sc-204774). Aclarubicin for in vivo mouse experiments was obtained from Shenzhen Main Luck Pharmaceuticals Inc. Aclarubicin (for in vitro experiments, sc-200160) and amrubicin (sc-207289) were obtained from Santa Cruz Biotechnology.

### Cell culture

K562, MM6, MOLM13, MV4:11, and U937 and THP-1 were cultured in RPMI-1640 medium supplemented with 8% fetal calf serum (FCS). OCI-AML2 and OCI-AML3 and OCI-AML4 were cultured in Iscove’s Modified Dulbecco’s medium (IMDM) supplemented with 8% FCS and glutamine. MelJuSo cells were maintained in IMDM supplemented with 8% FCS. All cell lines were maintained in a humidified atmosphere of 5% CO_2_ at 37 °C, regularly tested for the absence of mycoplasma and STR profile identified.

### Cell line construction

For endogenous tagged GFP-H2B K562 cells, mScarlet was swapped for GFP in the homology repair construct using NheI and BglII and cells were generated as described [5]. Co-transfection into K562 cells was done by electroporation using Lonza SF cell line kit. ABCB1 overexpressing K562 cells were generated as described [10]. Endogenous tagged 3×Flag-TopoIIα K562 cell line was generated using HR-3×Flag construct designed at least 40 base pairs up and downstream of the genomic TopoIIα stop codon. The gRNA target sequence was designed using the ZANG Lab CRISPR tool (http://crispr.mit.edu/) and cloned into the pX330 vector. Primers used for the HR construct: 5’-CACCGATGATCTGTTTTAAAATGTG-3’ and 5’-AAACCACATTTTAAAACAGATC ATC-3’. Co-transfection of ssDNA oligo and CRISPR plasmid (pX459) into K562 cells was performed by electroporation using Lonza SF cell line kit. Primers used for genotyping: forward primer: 5’-TAAGCAGAATTCATGCCACTTATTTGGGCAAT-3’ and reverse primer: 5’-TGCTTAAAGCTTTGCCCATGAGATGGTCACTA-3’.

### DNA damage assessed by Western blot and CFGE

After 2-hour drug treatment of indicated drugs at 5µM, cells were washed with PBS. WB and CFGE were performed as described [11]. Images were quantified with ImageJ.

### Fractionation assay

Endogenously tagged GFP-H2B cells (1×10^6^) were pre-incubated with protease inhibitor cocktail (P1860-1ML, Sigma-Aldrich) for 1 hour followed by 3-hour treatment with 10 µM of the indicated drugs. Cells were dissolved in lysis buffer (50mM Tris-HCl pH 8.0, 150 mM NaCl, 5 mM MgCl_2_, 0.5% NP40, 2.5% glycerol supplemented with protease inhibitors, 10 mM NMM) for 5 min on ice, and then centrifuged for 10 min at 15,000 g at 4°C. Both nucleus (pellet) and cytosol (supernatant) were washed once and submitted for Western Blot analysis. Primary antibodies used for detection: GFP (1:1000) [12], Lamin B1 (1:1000, 12987-I-AP, Proteintech), Calnexin (1:1000, C5C9, Cell signaling).

### Time-lapse confocal microscopy

For time-lapse confocal imaging, MelJuSo cells were seeded in 35-mm bottom petri dish (Poly-dlysin Coated, MatTek Corporation), transfected with TopoIIα-GFP construct (effectene, Qiagen) and imaged upon treatment with the indicated drugs [2]. Leica SP8 confocal microscope system, 63×lens, equipped with a climate chamber was used. TopoIIα-GFP distribution was quantified using Leica Application Suite X software.

### Short-term cell viability assay

Twenty-four hours after seeding into 96-well plates, cells were treated with indicated drugs for 2 hours at physiologically relevant concentrations [2]. Subsequently, drugs were removed by extensive washing, and cells were cultured for an additional 72 hours. Cell viability was measured using the CellTiter-Blue viability assay (Promega). Survival was normalized to the untreated control samples after correction for the background signal.

### ChIP-seq

Endogenous tagged 3×Flag-TopoIIα K562 cells were treated with 10 µM of indicated drugs for 4 hours. Cells were fixed and processed as described [3, 13]. ChIP was done with anti-Flag M2 (F3165, Sigma), followed by sequencing on an Illumina Hiseq2000 platform (Genome Sequencing Service Center of Stanford Center for Genomics and Personalized Medicine Sequencing Center).

ChIP-seq data were processed identically using the ENCODE Data Coordination Center (DCC) ChIP-seq pipeline (https://github.com/ENCODE-DCC/chip-seq-pipeline2) (v1.9.0). Briefly, the ChIP-seq reads were aligned to the human reference genome (GRCh37/hg19) using Bowtie2 [14]. Duplicate reads were removed using Picard MarkDuplicates (RRID:SCR_006525). Peaks of each sample were called against the whole-cell lysates replicates using SPP [15] with the parameters ‘-npeak 300000 -speak 155 -fdr 0.01’. The blacklisted regions described by ENCODE were discarded [16]. Reproducible peaks were intersected from two biological replicates and annotated with epigenomic signatures of K562, downloaded from the Roadmap Epigenomics Project [17]. Normalized TopoIIα binding affinity matrix: consensus peaks by samples, principle component analysis and differential binding affinity analysis were performed using the R package DiffBind [18].

### ATAC-seq

K562 cells were treated with 10 µM of indicated drugs for 4 hours. Cells were fixed and processed as described [19, 20]. DNA was processed using a customized library preparation method for ATAC-seq and was sequenced using an Illumina HiSeq4000 platform.

ATAC-seq data were processed identically using the ENCODE Data Coordination Center (DCC) ATAC-seq pipeline (https://github.com/ENCODE-DCC/atac-seq-pipeline) (v1.10.0). Briefly, the ATAC-seq reads were aligned to the human reference genome (GRCh37/hg19) using Bowtie2 [14]. Duplicate reads were removed using Picard MarkDuplicates (RRID:SCR_006525). Peaks were called using MACS2 [21] with the setting of ‘-p 0.01 -- shift -75 --extsize 150 --nomodel -B --SPMR --keep-dup all’, then followed by blacklisted regions filtering described by ENCODE [16]. Reproducible peaks were identified from two biological replicates and annotated with epigenomic signatures of K562, downloaded from the Roadmap Epigenomics Project [17]. ATAC-seq signal tracks for all the samples were generated by bedtools with the command ‘bedtools genomecov-scale’ using the read count per million (CPM) normalization and convert to bigwig files using bedGraphToBigWig. Normalized ATAC-seq read density matrix: consensus peaks by samples, principle component analysis and differential chromatin accessibility analysis were performed using the R package DiffBind [18].

### The bio-distribution of anthracyclines in mice

FVB/NRj mice ordered from Janvier Labs (Le Genest-Saint-Isle, France) were housed in individually ventilated cages under specific pathogen-free conditions in the animal facility of the NKI (Amsterdam, The Netherlands). All mouse experiments were approved by the Animal Ethics Committee of the NKI and were performed according to institutional and national guidelines. Male mice (8-week old) were i.v. injected with Doxo, Acla, Amr, Epi or Ida at 5 mg/kg (*n* = 5 per group). Four hours post injection, animals were sacrificed, and plasma, heart, lung, liver, kidney, spleen, brain, thymus, axillary and inguinal lymph nodes, and testis+epididymis were collected. Hearts were cut into two pieces with coronal section. One piece was fixed in EAF fixative (ethanol/acetic acid/formaldehyde/saline, 40:5:10:45 v/v/v/v) and processed for Phospho-H2AX (Ser139) IHC (1:100, #2577, Cell Signaling). The other half of the heart and the rest of organs were weighted, frozen and analyzed by LC-MS/MS [5].

### The cardiotoxicity of anthracyclines in mice

FVB/NRj mice (10–11-week old) were i.v. injected with 5 mg/kg of Doxo, 5 mg/kg of Acla, or 5 ml/kg of saline every 2 weeks for 4 times. After 4-week interval, the animals were i.v. injected with 5 mg/kg of indicated drug or 5 ml/kg of saline every 2 weeks for another 4 times. The mice were monitored every other day. When body weight loss was more than 20%, or circulation failure occurred, animal was euthanized by CO_2_. Subsequently, full body anatomy was performed. All organs were collected, fixed in EAF fixative and embedded in paraffin. Sections were cut at 2 μm from the paraffin blocks and stained with hematoxylin and eosin, and 4 μm for immunohistochemistry of Desmin (1:200, M0760, DakoCytomation), Vimentin (1:100, #5741, Cell Signaling), or Periostin (1:100, ab215199, Abcam). Pathology slides were reviewed twice by an expert mouse pathologist who was blind to the treatment. Incidence rate (IR = [number of mice with the specific side effect over a time period]/[sum of mice × time at risk during the same time period]) and cumulative incidence (CI = [number of mice with specific side effect at end time point]/[total number of mice at start]) were calculated for indicated side effects.

### r/rAML patients

Patients with refractory or relapse AML treated between July 2012 and July 2022 at Ruijin hospital, China were enrolled in this retrospective study. Some patients participated in trail ChiCTR-OPC-14005712, ChiCTR-OPC-15006896, ChiCTR-IIR-16008809, ChiCTR-OIC-16008952, ChiCTR-IIR-16008962 and ChiCTR-IIR-17011677. This study was approved by the ethics committee of Ruijin Hospital, all patients provided written informed consent. Cytogenetic risk was classified according to the modified Southwest Oncology Group criteria [22], and integrated risk was classified as described [23]. The treatment details were decribed in supplemental materials and methods. The baseline characteristics and clinical outcomes of the patients are summarized in supplemental Tables 1 and 2, respectively.

Cytogenetic risk was classified according to the modified Southwest Oncology Group criteria [22]: (1) favorable risk, including t(8;21) and inv(16) or t(16;16)(p13;q22); (2) unfavorable risk, including del(5q) or monosomy 5, monosomy 7 or del(7q), abnormal 3q, 9q, 11q, 21q, or 17p, t(6;9), t(9;22), and complex karyotypes (three or more unrelated chromosomes abnormal); and (3) intermediate risk, including normal karyotypes and all other anomalies. FLT3 internal tandem duplication and mutations in CEBPA, NPM1 and IDH1/2 were tested. Integrated risk was classified as described [23]. CR was defined as bone marrow blasts <5%, absolute neutrophil count ≥1×10^9^/L, and platelet count ≥100×10^9^/L, and absence of extramedullary disease. Partial remission was defined as having <15% (and a 50% decrease in bone marrow blasts) but >5% blasts or with <5% blasts but not reaching the CR criteria for blood cell count or clinical manifestation. The baseline characteristics and clinical outcomes of the patients are summarized in supplemental Tables 1 and 2, respectively.

### AML treatments

CAG patients were treated with 15LJ25 mg/m^2^ of Ara-C (cytarabine) injected s.c. every 12 h on days 1LJ14, 20 mg/d of Acla infused i.v. on days 1LJ4, and 200 μg/m^2^ of granulocyte stimulating factor (G-CSF) administered s.c. daily on days 1LJ14. G-CSF was reduced, or temporarily stopped when neutrophilia was >5×10^9^/L. IA patients were treated with 61.r10 mg/m^2^ of Ida infused i.v. on days 1LJ3 and 100LJ200 mg/m^2^ of Ara-C on days 1LJ7. VA patients were injected with 75 mg/m^2^ of azacitidine s.c. daily on days 1LJ7, and administered with venetoclax orally, once daily. The dose of venetoclax was 100 mg on day 1 and 200 mg on day 2; and 400 mg on days 3LJ28. In all subsequent 28-day cycles, the dose of venetoclax was initiated at 400 mg daily. The other induction chemotherapy for r/rAML patients included IA, DA, FLAG, CLAAG and CHA regimens. For patients treated with DA regimen, 20 mg/m^2^ of decitabine was administered i.v. daily on days 1LJ5, and 1 g/m^2^ of Ara-C was injected every 12 hours on days 6LJ7. For patients treated with FLAG regimen, fludarabine was infused i.v. at 30 mg/m^2^ on days 2LJ6; 4 hours after fludarabine infusion, Ara-C was injected i.v. at 1.51.r2 g/m^2^ over 3 hours on days 2LJ6; G-CSF was administered at 5 µg/kg s.c. on days 1LJ5; additional G-CSF may be administered since 7 days after the end of chemotherapy until WBC count >500/uL. For patients >60-year-old, the dose may be reduced to 20 mg/m^2^ for fludarabine and 0.5LJ1 g/m^2^ for cytarabine. For patients treated with CLAAG regimen, 5 mg/m^2^ of cladribine was infused i.v. over 2 hours on days 1LJ5; 15 mg/m^2^ of Ara-C was injected s.c. every 12 hours on days 1LJ10; trans retinoic acid (ATRA) was administered orally at 45 mg/m^2^ on days 4LJ6, then at 15 mg/m^2^ on days 7LJ20; 300 μg G-CSF was injected s.c. on day 0. For patients treated with CHA regimen, 5 mg/m^2^ of cladribine was infused i.v. over 2 hours on days 1LJ5; 2 mg/m^2^ of homoharringtonine was infused i.v. over 2 hours on days 1LJ5; 1 g/m^2^ of Ara-C was injected 2 hours after cladribine on days 1LJ5.

### Statistical analyses

Results are shown as mean ± SEM or mean ± SD. Statistical analysis was performed using Prism unless otherwise specified. All the in vitro experiments were performed with a minimum of three independent trials with the exception of ChIP-seq and ATAC-seq which were in biological duplicates. All the animals of mouse experiments are shown in the dot plots. Statistical tests are indicated in each figure legend.

## Results

### Mechanisms of action and cross-resistance of anthracyclines

To assess whether the various clinically administered anthracyclines (Fig. 1A) differ in their DNA- and chromatin-damaging activities, as well as cross-resistance, we exposed AML cell line THP1 at a clinically relevant dose for 2 hours followed by further culturing to mimic the pharmacokinetics in patients [24]. DNA damage was assessed by constant-field gel electrophoresis (CFGE) [11] and phosphorylation of H2AX at Ser139 (γH2AX) [25] as detected by Western blotting. Doxorubicin (Doxo), daunorubicin (Daun), epirubicin (Epi), idarubicin (Ida), and amrubicin (Amr) all induced DSBs, unlike aclarubicin (Acla) (Fig. 1B–E). Next, chromatin damage as the result of histone eviction was visualized by a fractionation assay using K562 cells with endogenously tagged GFP-H2B. Except for Amr, all the tested anthracyclines induced histone release from nucleus and accumulation in cytosol (Fig. 1F,G; Fig. S1A). Furthermore, all histone-evicting anthracyclines were able to redistribute GFP-TopoIIα on chromatin (Fig. S1B,C). Cytotoxicity assays revealed poor anti-cancer activity for the DNA-damaging analog Amr, while the other analogs bearing chromatin-damaging activity were effective in eliminating various myeloid leukemia cell lines (Fig. 1H).

**Figure 1.**
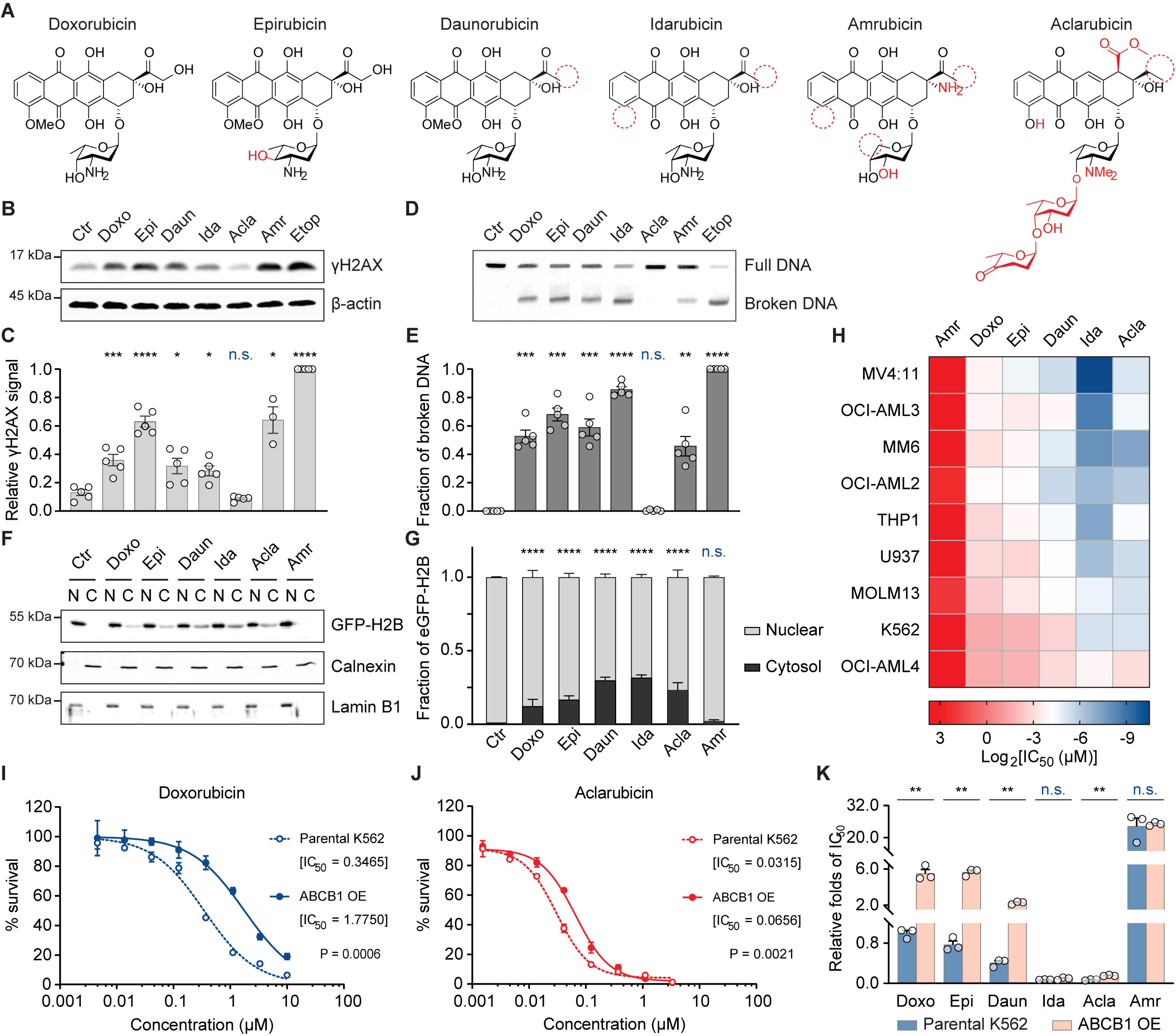
Acla differs from other anthracyclines in mechanisms of action and cross-resistance. (A) Structures of anthracyclines used in this study. Chemical moieties divergent from Doxo are depicted in red. (B) DNA damage examined by γH2AX Western blot. (C) Quantification of the γH2AX signal normalized to β-actin. Data are meanLJ±LJSEM; *n*LJ=LJ4 biological replicates; Student’s *t*-test. (D) DSBs analyzed by CFGE. (E) Quantification of relative broken DNA in (D). Data are meanLJ±LJSEM; *n*LJ=LJ4 biological replicates; Student’s *t*-test. (F) Histone eviction revealed by cell fractionation assay in K562-eGFP-H2B cells. N, nuclear fraction; C, cytosolic fraction. Calnexin and Lamin B1 are the loading control of each fraction. (G) The distribution of eGFP-H2B was quantified for both compartments. Data are mean ± SD; *n*LJ=LJ4 biological replicates; two-way ANOVA. (H) IC_50_ of each anthracycline in different leukemia cell lines. (I, J) Cell viability of parental K562 cells and ABCB1-overexpressing K562 cells upon Doxo (I) and Acla (J) treatment. Data are mean ± SD; *n*LJ=LJ3 biological replicates; two-way ANOVA. (K) Relative IC_50_ folds of each condition compared to that of Doxo in parental K562 cells. Data are meanLJ±LJSEM; *n*LJ=LJ3 biological replicates; Student’s *t*-test. \**P* < 0.05, \*\**P* < 0.01, \*\*\**P* < 0.001, and \*\*\*\**P* < 0.0001; n.s., not significant.

In addition to treatment-limiting cardiotoxicity, drug resistance could also limit the effect of anthracycline drugs. ABCB1 is a major drug efflux transporter contributing to anthracycline resistance [26]. We generated a Doxo-resistant leukemia cell line by overexpressing ABCB1 [10] and tested its sensitivity to other anthracyclines (Fig. 1I–K; Fig. S1D). The ABCB1-overexpressing cells acquired resistance to Doxo, Daun, Epi and Amr, but failed to export Ida and Acla efficiently (Fig. 1J,K). These data together indicate that clinically relevant anthracyclines should be considered as distinct drugs with unique features and different efficiencies. Evicting histones and relocalizing TopoIIα while not inducing DSBs, as in the case of Acla, apparently suffices for efficient cytotoxicity.

### Epigenetic selectivity of TopoIIα redistribution and histone eviction of anthracyclines

Anthracyclines poison TopoII by disrupting its interface with DNA by their sugar moieties and the cyclohexene ring [27]. However, the anthracycline-specific redistribution of TopoII and its association with histone eviction have not been characterized. We addressed this in K562 cells whose epigenomic information has been extensively profiled by ENCODE consortium [28]. Chromatin immunoprecipitation followed by deep sequencing (ChIP-seq) against endogenously tagged TopoIIα was performed 4 hours after anthracycline exposure in independent duplicate experiments (Fig. S2A). The resulting ChIP-seq profiles analyzed by principal component analysis (PCA) suggested that all histone-evicting anthracyclines cause extensive and drug-specific TopoIIα redistribution as compared to untreated or Amr-treated cells (Fig. 2A,B). To characterize this drug-specific TopoIIα redistribution, we coupled the ChIP-seq data to the epigenetic information of ENCODE (Fig. 2C). With the same sugar moiety and cyclohexane (Fig. 1A), Daun and Doxo (also labeled as hydroxyDaun) depleted TopoIIα from active chromatin regions, like DNase I hypersensitive regions (DHS), H3K4me1, H3K4me2, H3K4me3, H3K9ac, H3K27ac, H3K79me2 and H2A.Z [29]. Instead, TopoIIα was trapped in compact chromatin regions marked by H3K27me3 [29], H3K9me1 [30] and H3K9me3 [31] (Fig. 2C). Ida and Acla depleted TopoIIα from broader chromatin states, except for H3K9me3 [31], H3K27me3 [29] and H3K36me3 [31] decorated regions (Fig. 2C). Epi, with an epimerizing hydroxyl group at the sugar moiety (Fig. 1A), further depleted TopoIIα from H3K36me3 [31] modified regions (Fig. 2C). It is worth noting that the redistribution of TopoIIα at regions with unknown epigenetic features was increased for all drugs, except for Amr (Fig. 2C).

**Figure 2.**
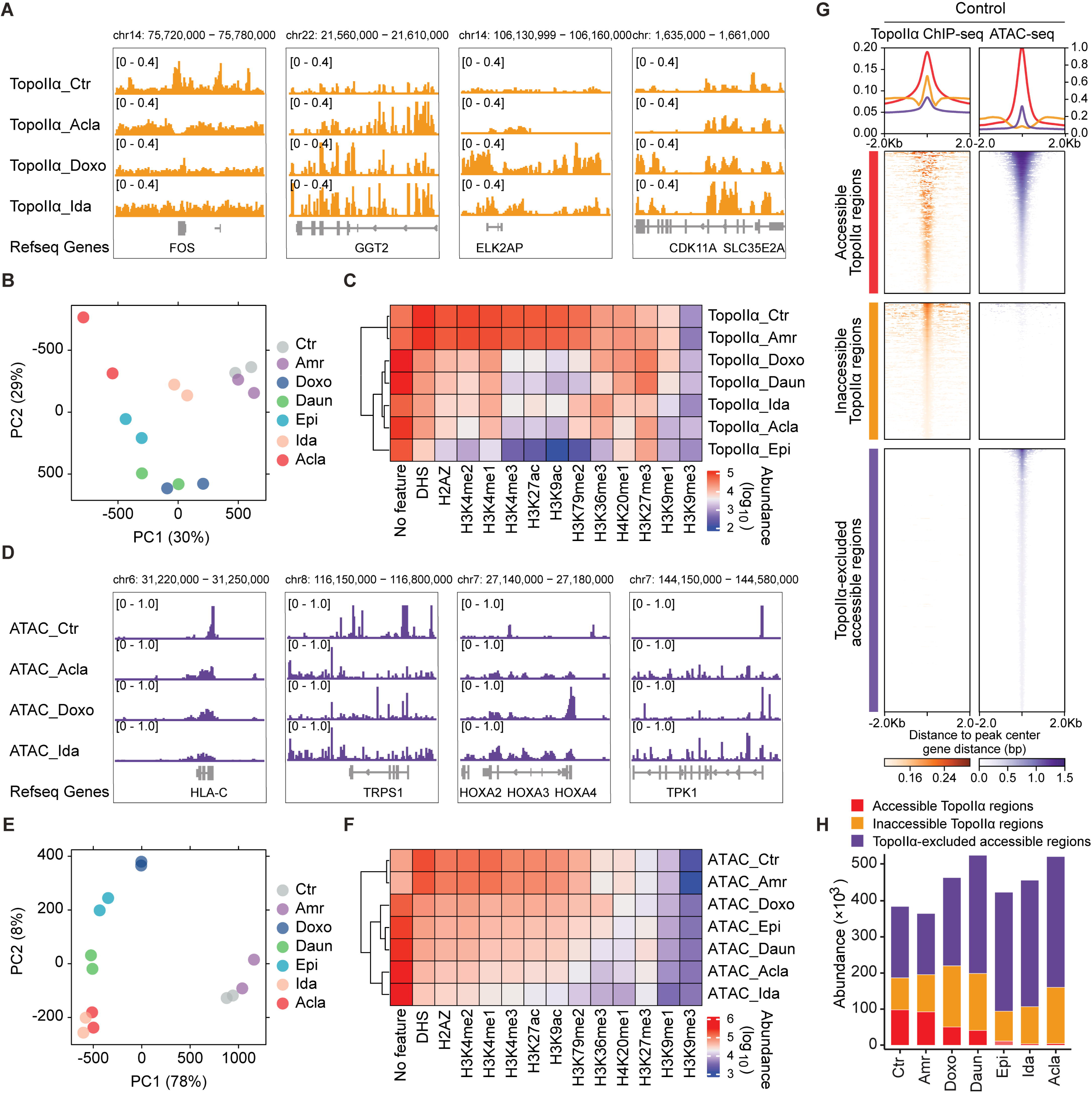
Epigenetic selectivity of TopoIIα redistribution and histone eviction of anthracyclines. (A) Illustration of drug-specific TopoIIα redistribution revealed by ChIP-seq. (B) Principal component analysis (PCA) of TopoIIα ChIP-seq data. Two independent biological replicates were included for each condition. (C) Heatmap of TopoIIα peak abundance associated with specific histone features derived from Roadmap Epigenomics Project. (D) Illustration of drug-specific accessible chromatin regions revealed by ATAC-seq. (E) PCA analysis of ATAC-seq data. Two independent biological replicates were included for each condition. (F) Heatmap of ATAC peak abundance associated with specific histone. (G) Density plots showing the accessible and inaccessible TopoIIα regions, and TopoIIα-excluded accessible regions surrounding ±2 Lkb from the center of the detected peaks in the untreated K562 cells. (H) The abundance of each category in different conditions.

While the TopoIIα redistribution showed drug specificity, we continued to identify histone eviction preferences of each anthracycline using transposase-accessible chromatin with sequencing (ATAC-seq) [19] in K562 cells (Fig. 2D). In line with its lack of chromatin-damaging activity (Fig. 1F,G), Amr failed to induce *de novo* open chromatin and could therefore be found close to untreated cells in PCA plot (Fig. 2E). Similar to the TopoIIα redistribution pattern, the other tested anthracyclines evicted histones mostly from open chromatin regions, once again exhibiting distinct preferences (Fig. 2F). As histone modifications are required for recruitment of TopoIIα to DNA [32], selective eviction of histones from open chromatin regions may contribute to the observed depletion of TopoIIα from the same regions. Based on the comparative analysis of TopoIIα ChIP-seq and the ATAC-seq experiments, we observed that regions identified by both ATAC-seq and TopoIIα ChIP-seq were greatly decreased by histone-evicting anthracyclines (Fig. 2G,H; Fig. S2B). Nonetheless, outside of overlapping regions, TopoIIα redistribution and *de novo* accessible chromatin were usually not associated with each other, indicating that these may constitute two independent effects of anthracyclines (Fig. S2B). Thus, integration of TopoIIα ChIP-seq and ATAC-seq revealed the connection between histone eviction and TopoIIα depletion in transcriptionally active regions, as well as selective impact of each analog on the epigenetic landscape. Each clinically used anthracycline has individual genomic preferences, which may define drugs with different specificity.

### Bio-distribution of anthracyclines

Acla is effective in eliminating a large variety of cancer cells in tissue culture [5]. However, it is mainly used in treating AML and is less effective against solid tumors [33, 34]. This could be the result of differences in biodistribution of anthracyclines. To test this, we performed a comprehensive pharmacokinetic study for Acla and the other clinically used anthracyclines in mice. Four hours post iv injection of a clinically relevant dose of indicated drug at 5 mg/kg [35], Doxo, Epi and Ida showed similar bio-distribution patterns across different organs (Fig. 3A; Fig. S3A). Amr had a similar tissue distribution as Doxo, but showed reduced distribution to lung, kidneys, and heart. Brain tissue was poorly penetrated by all drugs (Fig. 3A; Fig. S3A). A notable exception was Acla, which accumulated in lymphoid organs (spleen, thymus and lymph nodes) but poorly distributed to other organs (Fig. 3A; Fig. S3A). This might be an explanation why Acla is ineffective in treating solid tumors, as opposed to hematologic tumors.

**Figure 3.**
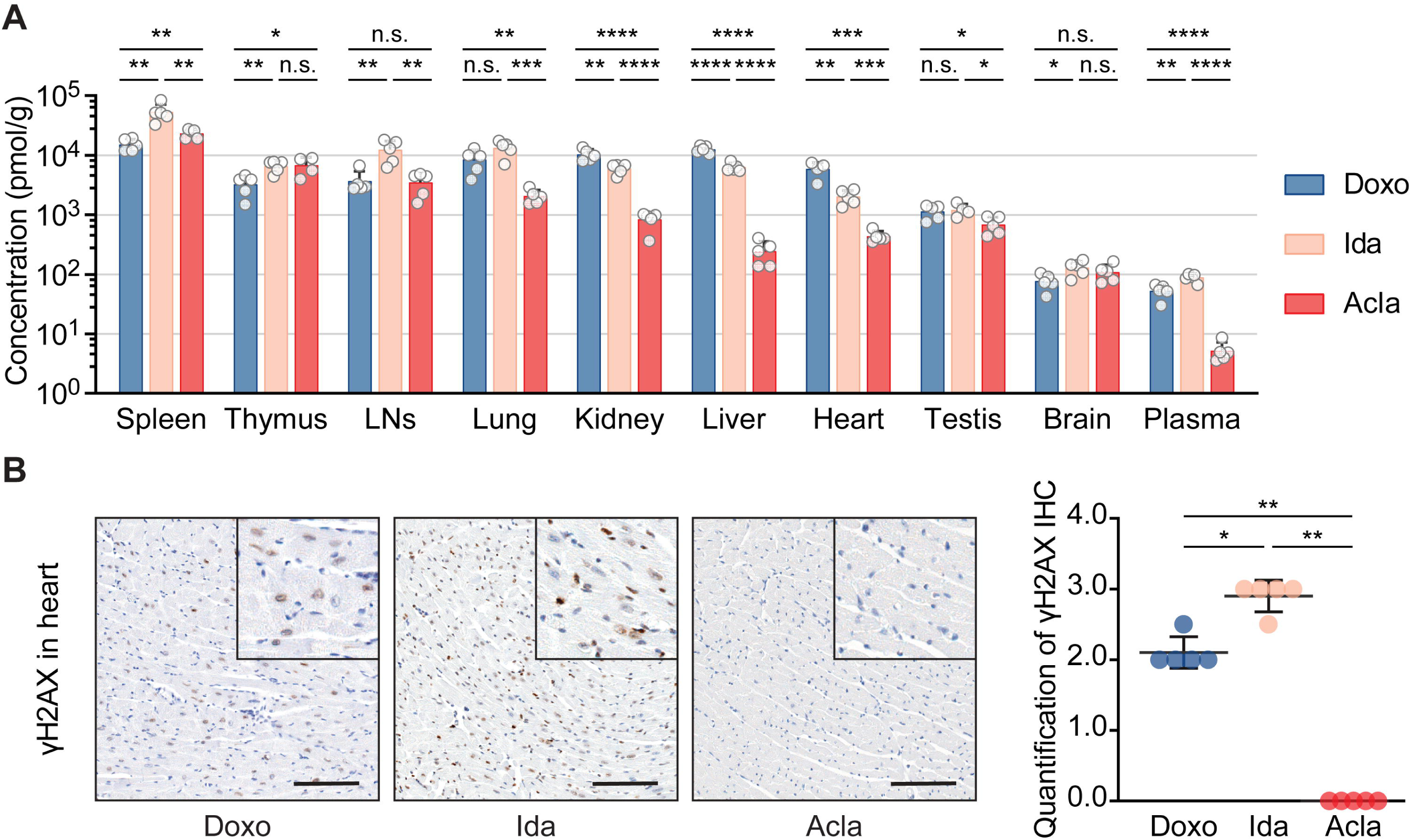
Bio-distribution of clinically-used anthracyclines in mice. (A) Drug bio-distribution was determined 4 hours after i.v. injection of indicated drug at 5 mg/kg. Data are represented as meanL± SD from 5 mice per group. Student’s *t*-test. (B) Representative microscopic images of γH2AX IHC staining of the hearts. Scale bars, 100 μm. Quantification is represented as meanL±LSD, *n* = 5, Mann-Whitney test. \**P* < 0.05, \*\**P* < 0.01, \*\*\**P* < 0.001, and \*\*\*\**P* < 0.0001; n.s., not significant.

We further evaluated DNA damage in the heart in response to drug exposure. While Doxo, Epi and Ida incited persistent DNA damage in the heart, the hearts of Acla- and Amr-treated mice did not show any γH2AX signals 4 hours after drug administration (Fig. 3B; Fig. S3B), in line with poor tissue penetration of Acla and Amr and the fact that Acla does not induce DNA damage at all.

### Acla is safe and well tolerated following Doxo treatment

Doxo, Daun, Ida and Epi all induce cumulative dose-dependent cardiotoxicity, which limits patient treatment to only few courses. Since Acla treatment is not associated with cardiotoxicity [5], we reasoned that it could be safe to apply this drug to mice following treatment with cardiotoxic anthracyclines. To test this, Acla was administered to mice after half-maximum cumulative dose of Doxo (Fig. 4A) and followed over time. Cardiotoxicity induced by Doxo caused cardiac hypertrophy and remodeling that presented as thrombus formation in the left atrium and auricle of the heart accompanied by inflammation, fibrosis and calcification [5, 36] (Fig. 4B; Fig. S4A). Further staining for profibrotic proteins vimentin [37] and periostin [38], and cytoskeletal protein desmin [39], revealed impairment of myocytes and fibrosis in the stroma (Fig. 4C–H; Fig. S4B–F). With impaired cardiac function, Doxo-treated mice died from circulation failure (Fig. 4I).

**Figure 4.**
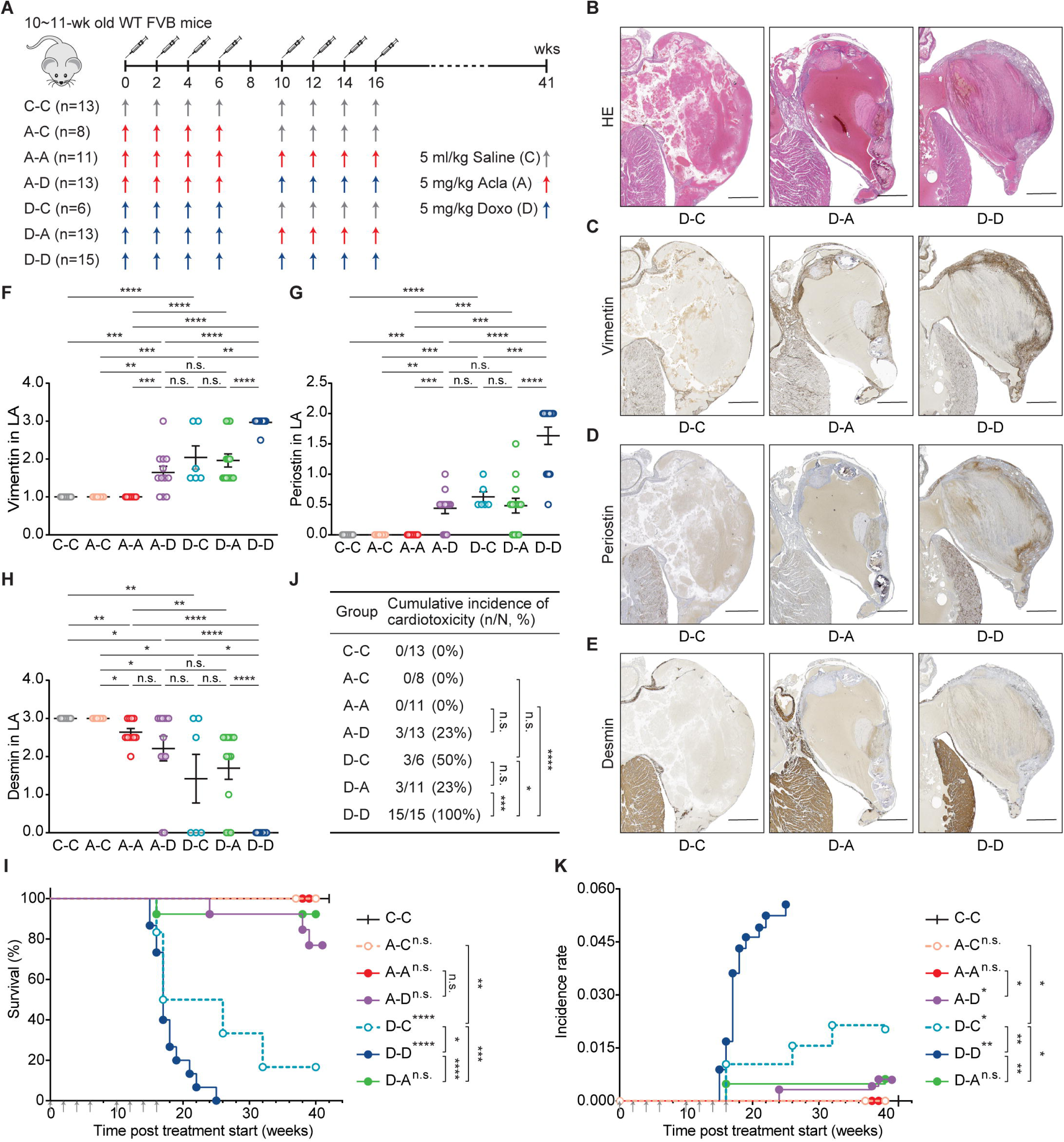
Acla is safe and well tolerated following Doxo treatment. (A) Wild-type FVB mice were i.v. injected with Doxo (D), Acla (A), or saline (C) as indicated. (BLE) Representative microscopic images of the left atrium (LA) of heart. Lesions caused by Doxo treatment represent as impairment of the wall, thrombosis, inflammation, fibrosis/calcification, and disappearing of the lumen of atrium (filled by a large thrombus). Scale bars, 500 μm. (FLH) Quantification of the indicated IHC staining in LA. Data are meanL±LSEM, Mann-Whitney test. (I) Animal survival is plotted in Kaplan-Meier curves. Log-rank test. (J) Cumulative incidence of cardiotoxicity. Fisher’s exact test. (K) Incidence rate of cardiotoxicity. Two-way ANOVA with repeated measures, two-sided. \**P* < 0.05, \*\**P* < 0.01, \*\*\**P* < 0.001, and \*\*\*\**P* < 0.0001; n.s., not significant.

Reflecting clinical observations [40] and our prior work [5], the incidence, latency and histopathological alterations of Doxo-induced cardiotoxicity developed in a dose-dependent manner (Fig. 4A, D-C vs D-D), whereas eight courses of Acla treatment showed no abnormalities in the heart (Fig. 4F–K; Fig. S4, A-A vs C-C/D-C/D-D). More importantly, Acla treatment after half-maximum cumulative dose of Doxo did not aggravate cardiotoxicity (Fig. 4; Fig. S4, D-A vs D-D). Similarly, pre-treatment with Acla did not make mice more susceptible to Doxo-induced cardiotoxicity (Fig. 4; Fig. S4, A-D vs D-C/D-D). Hence, Acla is safe and well tolerated following Doxo treatment in this model system. Considering limited cardiotoxicity, little cross-resistance and differences in epigenomic specificities between anthracyclines, Acla would be predicted to act as an independent drug in salvage relapsed/refractory AML (r/rAML) patients who have already received the maximum cumulative dose of cardiotoxic anthracyclines. Such systematic studies have not been reported.

### CAG strongly improves the survival of relapsed/refractory AML patients

R/rAML is one of the most challenging situations in hematology, with a 5-year overall survival (OS) of only 10% [41]. The prognosis of elderly or unfit r/rAML patients is even more disappointing [42]. Unfortunately, no specific salvage regimen has appeared as a standard treatment for r/rAML [42]. CAG regimen, containing low-dose of cytarabine, Acla and G-CSF, has been widely used in China and Japan for treating AML [43, 44]. It is well tolerated by r/r and elderly AML patients with lower toxicity than conventional chemotherapies [43–45]. To compare the efficacy of CAG to other salvage chemotherapies in r/rAML, we conducted a single-center retrospective study at Ruijin hospital (Shanghai, China).

A total of 186 r/rAML patients, treated between July 2012 and July 2022, were included in the study. In this group of patients, fit patients with *de novo* AML often received Ida in combination with cytarabine (IA) rather than Daun or Doxo for first-line therapy, as Ida is more effective for induction therapy in AML [46, 47]. However, all three anthracyclines are cardiotoxic [48]. Among the patients who developed refractory and relapse after primary IA treatment for *de novo* disease, 66 patients received CAG regimen (2^nd^-line CAG group), 67 received other chemotherapy regimens (2^nd-^line others group), and 34 received emerging target therapy regimen venetoclax + azacitidine (2^nd^-line VA group) (Fig. 5A). 19 patients who were treated with CAG for both *de novo* and r/r diseases were enrolled to illustrate that Acla does not contribute to cardiotoxicity and is compatible with extended treatment (1^st^&2^nd^-line CAG group) (Fig. 5A). Notably, at Ruijin hospital, CAG is usually applied to *de novo* AML patients with unfit conditions who are not expected to tolerate conventional intensive chemotherapies. The demographic and clinical characteristics of all patients at r/rAML diagnosis are summarized in supplemental Table 1. There was no significant difference between 2^nd^-line CAG group and any other group with respect to age, gender, French-American-British type, cytogenetic risk, and frequent oncogenic mutations. Slightly more patients in the 2^nd^-line others group were diagnosed with favorable integrated risk than in the other arms of the study.

**Figure 5.**
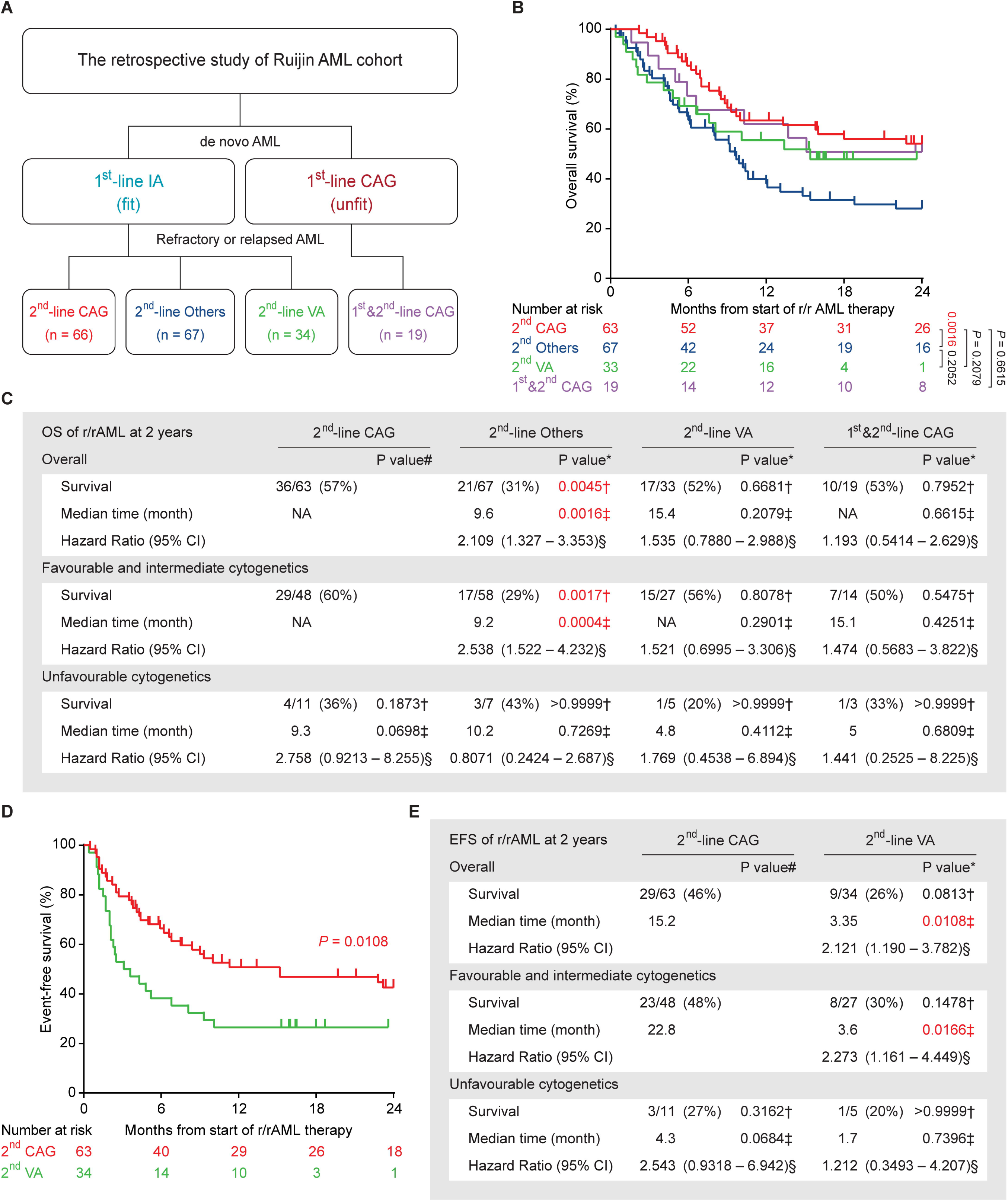
CAG is effective in treating r/rAML patients. (A) The therapy overview of Ruijin AML cohort. CAG: cytarabin, aclarubicin, G-CSF; IA: idarubicin, cytarabine; VA: venetoclax, azacitidine; Others: chemotherapy regimens other than CAG. (B) Overall survival of the r/rAML patients from the start of r/rAML induction treatment to the date of death. Log-rank test. (C) The statistical analysis of OS of r/rAML patients at 2 years. Data are n/N (%), * compared with 2^nd^-line CAG group, † Fisher’s exact test, ‡ Log-rank test, § Mantel-Haenszel test, # compared with 2^nd^-line CAG patients with favorable and intermediate cytogenetics. (D) Event-free survival of the r/rAML patients from the start of r/rAML induction treatment to the date of relapse/refractory/death. (E) The statistical analysis of EFS of r/rAML patients at 2 years. Data are n/N (%), * compared with 2^nd^-line CAG group, † Fisher’s exact test, ‡ Log-rank test, § Mantel-Haenszel test, # compared with 2^nd^-line CAG patients with favorable and intermediate cytogenetics.

The overall complete remission (CR) rate after induction was 74% for the 2^nd^-line CAG group, which is significantly higher than that of the 2^nd^-line others (37%) and the 2^nd^-line VA group (35%) (Table 1). Patients with favorable and intermediate cytogenetics or integrated risk responded better to CAG. However, cytogenetics or integrated risk was not prognostic for the other groups (Table 1). Among the patients with a favorable and intermediate risk, the complete remission rate of 2^nd^-line CAG was superior to that of 2^nd^-line others and the 2^nd^-line VA group (Table 1). While age is a poor prognostic factor for AML [42], no significant difference was seen for CAG outcome in patients <60- and ≥60-year-old. Actually, for patients ≥60-year-old, 2^nd^-line CAG also outperformed other treatment regimens (Table S2). No CR and major prognostic mutation associations were detected for any treatment group due to small cohort (Table S2). In terms of CR rate, compared to the 2^nd^-line CAG group that first received IA, the 1^st^&2^nd^-line CAG group performed slightly less optimally due to unfit conditions. However, this treatment protocol was not inferior to the 2^nd^-line others and the 2^nd^-line VA group (Table 1; Table S2), suggesting that CAG remains effective and tolerable in unfit r/rAML patients, even after prior CAG treatments.

**Table 1.**
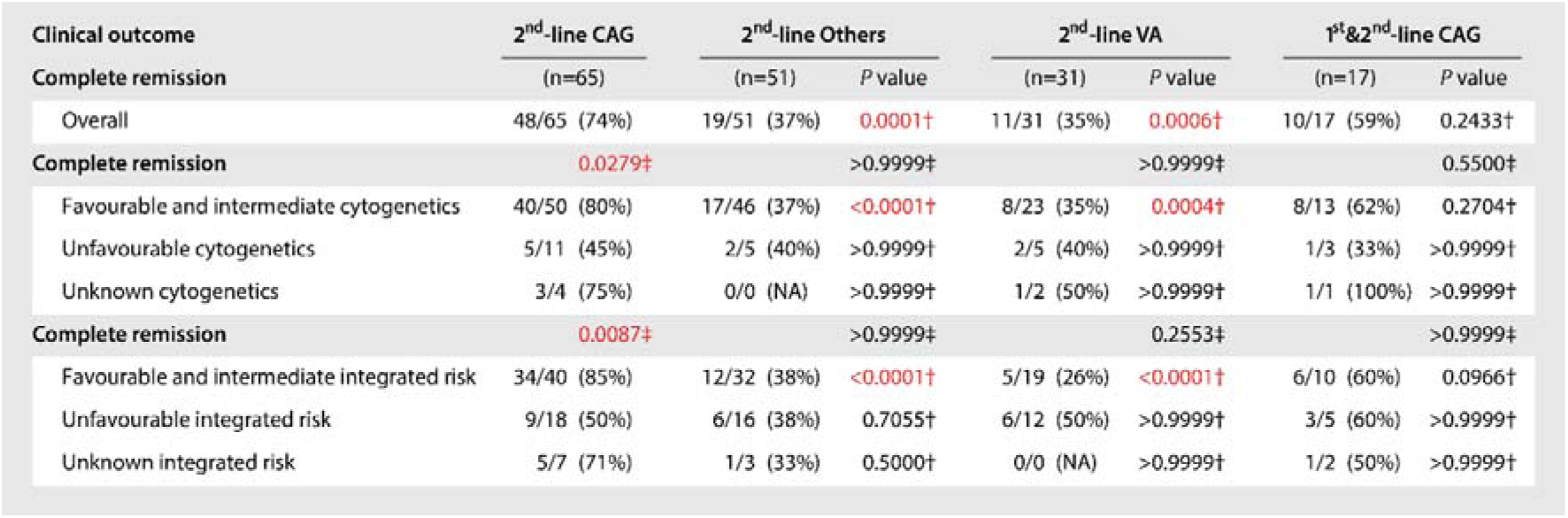
Treatment outcomes of r/rAML patients. CAG: cytarabin, aclarubicin, G-CSF; IA: idarubicin, cytarabine; VA: venetoclax, azacitidine; Others: chemotherapy regimens other than CAG. Data are n/N (%). † Fisher’s exact test between the indicated group and 2^nd^-line CAG group. ‡ Fisher’s exact test within group.

| Clinical outcome | 2<sup>nd</sup>-line CAG | 2<sup>nd</sup>-line Others |  | 2<sup>nd</sup>-line VA |  | 1<sup>st</sup>&2<sup>nd</sup>-line CAG |  |
| --- | --- | --- | --- | --- | --- | --- | --- |
| Complete remission | (n=65) | (n=51) | P value | (n=31) | P value | (n=17) | P value |
| Overall | 48/65 (74%) | 19/51 (37%) | 0.0001† | 11/31 (35%) | 0.0006† | 10/17 (59%) | 0.2433† |
| Complete remission | 0.0279‡ |  | >0.9999‡ |  | >0.9999‡ |  | 0.5500‡ |
| Favourable and intermediate cytogenetics | 40/50 (80%) | 17/46 (37%) | <0.0001† | 8/23 (35%) | 0.0004† | 8/13 (62%) | 0.2704† |
| Unfavourable cytogenetics | 5/11 (45%) | 2/5 (40%) | >0.9999† | 2/5 (40%) | >0.9999† | 1/3 (33%) | >0.9999† |
| Unknown cytogenetics | 3/4 (75%) | 0/0 (NA) | >0.9999† | 1/2 (50%) | >0.9999† | 1/1 (100%) | >0.9999† |
| Complete remission | 0.0087‡ |  | >0.9999‡ |  | 0.2553‡ |  | >0.9999‡ |
| Favourable and intermediate integrated risk | 34/40 (85%) | 12/32 (38%) | <0.0001† | 5/19 (26%) | <0.0001† | 6/10 (60%) | 0.0966† |
| Unfavourable integrated risk | 9/18 (50%) | 6/16 (38%) | 0.7055† | 6/12 (50%) | >0.9999† | 3/5 (60%) | >0.9999† |
| Unknown integrated risk | 5/7 (71%) | 1/3 (33%) | 0.5000† | 0/0 (NA) | >0.9999† | 1/2 (50%) | >0.9999† |

Since both CAG and VA therapy were well tolerated, some patients continued with these regimens for consolidation therapy and long-term maintenance. Among these were patients treated for up to 11 times with CAG without any cardiac issues, far more than other anthracyclines (Fig. S5A). This strategy was not an option for IA treatment due to accumulated cardiotoxicity and resistance. Superior CR rates and/or sustainable treatment schedule of CAG and VA translated into a more than 20% better OS as compared to 2^nd^-line others group (Fig. 5B,C; Fig. S5B,C). Nonetheless, event-free survival analysis indicated a better durable response of 2^nd^-line CAG when compared to 2^nd^-line VA (Fig. 5D,E), possibly due to the development of VA resistance [49]. Of note, the 1^st^&2^nd^-line CAG patients were selected for the limited toxicity of this regimen due to unfit conditions, which is associated with poor prognosis. Still, these patients achieved comparable OS as the 2^nd^-line CAG group (Fig. 5B,C; Fig. S5D,E). Taken together, these findings reveal that CAG regimen can be a superior, low-toxicity chemotherapy for r/rAML, including those with unfit conditions.

## Discussion

Despite their devastating side effect profiles, anthracyclines including doxorubicin, daunorubicin and idarubicin have been cornerstones of oncology treatment for over 50 years. While the molecular mechanisms of action have been mostly considered the same across different anthracycline analogs, clinical observations and our previous studies have indicated that this does not apply to all variants [2, 3, 5, 50]. Here, we performed a comprehensive analysis of clinically relevant anthracyclines against key parameters including mode of action, epigenomic selectivity and clinical performance, and find individual anthracyclines have substantially different pharmacological properties. In the course of our evaluation, Acla stood out as it is minimally cardiotoxic and prefers lymphoid organs over solid tissues. Attributing to its high therapeutic index for AML, we demonstrate that Acla can be safely used in second-line therapy after initial treatment of cardiotoxic anthracyclines, leading to 23% higher 5-year OS for r/rAML patients.

A common feature of anthracyclines is their poisoning of TopoIIα. TopoIIα plays a role in all areas of chromosome structure, from nucleosome assembly and disassembly to chromosome condensation and segregation between daughter cells [51–53]. DSB induction mediated by the catalytic activity of TopoIIα has always been considered as the primary cytotoxic mechanism of anthracyclines [1]. However, this dogma is challenged by Acla and other anthracycline variants whose major cytotoxic activity is chromatin damage, also known as histone eviction [54]. It has been reported that deregulation of TopoIIα leads to cell death in a manner independent of its catalytic activity [55, 56]. Therefore, without poisoning TopoIIα for DNA damage, depleting and redistributing TopoIIα are already detrimental, as illustrated by Acla. This finding further uncovers the anti-cancer mechanism of chromatin damage caused by histone eviction.

While Acla was first introduced into clinic 40 years ago (it was available in Europe until 2004 and was never registered in the U.S.), it is surprising that the unique effects of Acla in r/rAML has never been realized. Our data suggest that applying CAG therapy in r/rAML patients would increase their 5-year OS by 23%. If Acla was incorporated into current AML regimens, 10,000 r/rAML patients in the U.S. and Europe [57] would survive annually from this off-patent and low-cost drug. The impact on pediatric oncology will be even more profound, as 50% of childhood cancer patients receive high-dose anthracyclines and go on to have 5- to 15-fold increased risk for heart failure compared to the general population [9]. Introducing Acla in their treatment would mitigate this side effect. Considering old drugs in new ways may therefore substantially – and expeditiously – improve the survival and quality of life of many cancer patients, as exemplified here.

## Data Availability

All data produced in the present study are available upon reasonable request to the authors and with permission of Ruijin hospital.

https://www.ncbi.nlm.nih.gov/geo/query/acc.cgi?acc=GSE240443

## Abbreviations

Acla: Aclarubicin
AML: Acute myeloid leukemia
Amr: Amrubicin
Ara-C: Cytarabine
ATAC-seq: Assay for Transposase-Accessible Chromatin using sequencing
ATRA: Retinoic acid
CAG: cytarabine, Acla and G-CSF
CFGE: Constant-field gel electrophoresis
ChIP-seq: Chromatin immunoprecipitation followed by sequencing
CR: Complete remission
Daun: Daunorubicin
DHS: DNase I hypersensitive regions
Doxo: Doxorubicin
DSBs: DNA double-strand breaks
Epi: Epirubicin
FCS: Fetal calf serum
G-CSF: Granulocyte colony stimulating factor
GEO: Gene expression omnibus
IA: Ida in combination with cytarabine
Ida: Idarubicin
IMDM: Iscove’s modified dulbecco’s medium
IR: Incidence rate
OS: Overall survival
PCA: Principal component analysis
*γ*H2AX: phosphorylation of H2AX at Ser139
r/rAML: relapsed/refractory AML
SDS: Sodium dodecyl sulfate
TopoII: Topoisomerase II

## Supplementary Information

Supplementary Materials.dox includes Figures S1 to S5 and Tables S1 to S2.

## Declarations

### Ethics approval and consent to participate

All mouse experiments were approved by the Animal Ethics Committee of the NKI and were performed according to institutional and national guidelines. The retrospective clinical study was approved by the ethics committee of Ruijin Hospital, and all patients provided written informed consent.

### Availability of data and materials

The sequencing data supporting the findings of this study are available at the Gene Expression Omnibus (GEO) under accession numbers GSE240443 (https://www.ncbi.nlm.nih.gov/geo/query/acc.cgi?acc=GSE240443). Source data are provided with this paper. Clinical data are available from the corresponding authors upon reasonable request and with permission of Ruijin hospital.

### Authors’ contribution

Contribution: X.Q., S.Y.v.d.Z. and J.N. designed experiments. X.Q. and S.Y.v.d.Z. performed experiments. O.v.T., F.L.H., and M.A.v.G. contributed to experiments. Y.Z. and X.L. collected the r/rAML data. X.Q., X.L., J.L. and J.N. analyzed the clinical data. M.T. and B.P. performed and analyzed ChIP-seq and ATAC-seq experiments. L.J., Y.L. and B.P. made stable cell lines. J.S. performed pathological analyses. C.L.Z. contributed to the result interpretation. X.Q., S.Y.v.d.Z. and J.N. wrote the manuscript, with the input of all authors. All authors read and approved the final manuscript.

## Acknowledgements

The authors thank the people from the Preclinical Intervention Unit of the Mouse Clinic for Cancer and Ageing (MCCA) at NKI for their technical support performing the animal experiments and the staff from the Experimental Animal Pathology facility at NKI for their service on histochemistry and immunochemistry staining; Prof. J.P. Vandenbroucke and M. Schaapveld for help with the epidemiological assessment of clinical data; J. Sarthy and I. Berlin for comments and critical reading of the Manuscript.

## References

1. Tewey KM, Rowe TC, Yang L, Halligan BD, Liu LF. Adriamycin-induced DNA damage mediated by mammalian DNA topoisomerase II. Science. 1984;226(4673):466–8.

2. Pang B, Qiao X, Janssen L, Velds A, Groothuis T, Kerkhoven R, et al. Drug-induced histone eviction from open chromatin contributes to the chemotherapeutic effects of doxorubicin. Nature communications. 2013;4:1908.

3. Pang B, de Jong J, Qiao X, Wessels LF, Neefjes J. Chemical profiling of the genome with anti-cancer drugs defines target specificities. Nature chemical biology. 2015;11(7):472–80.

4. Yang F, Kemp CJ, Henikoff S. Doxorubicin enhances nucleosome turnover around promoters. Curr Biol. 2013;23(9):782–7.

5. Qiao X, van der Zanden SY, Wander DPA, Borras DM, Song JY, Li X, et al. Uncoupling DNA damage from chromatin damage to detoxify doxorubicin. Proceedings of the National Academy of Sciences of the United States of America. 2020;117(26):15182–92.

6. Girling DJ. Comparison of oral etoposide and standard intravenous multidrug chemotherapy for small-cell lung cancer: a stopped multicentre randomised trial. Medical Research Council Lung Cancer Working Party. Lancet. 1996;348(9027):563–6.

7. Hong WK, Nicaise C, Lawson R, Maroun JA, Comis R, Speer J, et al. Etoposide combined with cyclophosphamide plus vincristine compared with doxorubicin plus cyclophosphamide plus vincristine and with high-dose cyclophosphamide plus vincristine in the treatment of small-cell carcinoma of the lung: a randomized trial of the Bristol Lung Cancer Study Group. Journal of Clinical Oncology. 1989;7(4):450–6.

8. Sekeres MA, Guyatt G, Abel G, Alibhai S, Altman JK, Buckstein R, et al. American Society of Hematology 2020 guidelines for treating newly diagnosed acute myeloid leukemia in older adults. Blood Adv. 2020;4(15):3528–49.

9. Armenian S, Bhatia S. Predicting and Preventing Anthracycline-Related Cardiotoxicity. Am Soc Clin Oncol Educ Book. 2018;38:3–12.

10. Li Y, Tan M, Sun S, Stea E, Pang B. Targeted CRISPR activation and knockout screenings identify novel doxorubicin transporters. Cell Oncol (Dordr). 2023.

11. Wlodek D, Banath J, Olive PL. Comparison between pulsed-field and constant-field gel electrophoresis for measurement of DNA double-strand breaks in irradiated Chinese hamster ovary cells. International journal of radiation biology. 1991;60(5):779–90.

12. van der Kant R, Fish A, Janssen L, Janssen H, Krom S, Ho N, et al. Late endosomal transport and tethering are coupled processes controlled by RILP and the cholesterol sensor ORP1L. J Cell Sci. 2013;126(Pt 15):3462–74.

13. Schmidt D, Wilson MD, Spyrou C, Brown GD, Hadfield J, Odom DT. ChIP-seq: using high-throughput sequencing to discover protein-DNA interactions. Methods. 2009;48(3):240–8.

14. Langmead B, Salzberg SL. Fast gapped-read alignment with Bowtie 2. Nat Methods. 2012;9(4):357–9.

15. Kharchenko PV, Tolstorukov MY, Park PJ. Design and analysis of ChIP-seq experiments for DNA-binding proteins. Nat Biotechnol. 2008;26(12):1351–9.

16. Amemiya HM, Kundaje A, Boyle AP. The ENCODE Blacklist: Identification of Problematic Regions of the Genome. Scientific reports. 2019;9(1):9354.

17. Roadmap Epigenomics C, Kundaje A, Meuleman W, Ernst J, Bilenky M, Yen A, et al. Integrative analysis of 111 reference human epigenomes. Nature. 2015;518(7539):317–30.

18. Ross-Innes CS, Stark R, Teschendorff AE, Holmes KA, Ali HR, Dunning MJ, et al. Differential oestrogen receptor binding is associated with clinical outcome in breast cancer. Nature. 2012;48.

19. Buenrostro JD, Giresi PG, Zaba LC, Chang HY, Greenleaf WJ. Transposition of native chromatin for fast and sensitive epigenomic profiling of open chromatin, DNA-binding proteins and nucleosome position. Nat Methods. 2013;10(12):1213–8.

20. Corces MR, Trevino AE, Hamilton EG, Greenside PG, Sinnott-Armstrong NA, Vesuna S, et al. An improved ATAC-seq protocol reduces background and enables interrogation of frozen tissues. Nat Methods. 2017;14(10):959–62.

21. Zhang Y, Liu T, Meyer CA, Eeckhoute J, Johnson DS, Bernstein BE, et al. Model-based analysis of ChIP-Seq (MACS). Genome biology. 2008;9(9):R137.

22. Slovak ML, Kopecky KJ, Cassileth PA, Harrington DH, Theil KS, Mohamed A, et al. Karyotypic analysis predicts outcome of preremission and postremission therapy in adult acute myeloid leukemia: a Southwest Oncology Group/Eastern Cooperative Oncology Group Study. Blood. 2000;96(13):4075–83.

23. Dohner H, Estey EH, Amadori S, Appelbaum FR, Buchner T, Burnett AK, et al. Diagnosis and management of acute myeloid leukemia in adults: recommendations from an international expert panel, on behalf of the European LeukemiaNet. Blood. 2010;115(3):453–74.

24. Greene RF, Collins JM, Jenkins JF, Speyer JL, Myers CE. Plasma pharmacokinetics of adriamycin and adriamycinol: implications for the design of in vitro experiments and treatment protocols. Cancer research. 1983;43(7):3417–21.

25. Kuo LJ, Yang LX. Gamma-H2AX - a novel biomarker for DNA double-strand breaks. In vivo. 2008;22(3):305–9.

26. Cox J, Weinman S. Mechanisms of doxorubicin resistance in hepatocellular carcinoma. Hepat Oncol. 2016;3(1):57–9.

27. Wang AH, Ughetto G, Quigley GJ, Rich A. Interactions between an anthracycline antibiotic and DNA: molecular structure of daunomycin complexed to d(CpGpTpApCpG) at 1.2-A resolution. Biochemistry. 1987;26(4):1152–63.

28. Gerstein MB, Kundaje A, Hariharan M, Landt SG, Yan KK, Cheng C, et al. Architecture of the human regulatory network derived from ENCODE data. Nature. 2012;489(7414):91–100.

29. Barski A, Cuddapah S, Cui K, Roh TY, Schones DE, Wang Z, et al. High-resolution profiling of histone methylations in the human genome. Cell. 2007;129(4):823–37.

30. Alvarez F, Munoz F, Schilcher P, Imhof A, Almouzni G, Loyola A. Sequential establishment of marks on soluble histones H3 and H4. The Journal of biological chemistry. 2011;286(20):17714–21.

31. Rea S, Eisenhaber F, O’Carroll D, Strahl BD, Sun ZW, Schmid M, et al. Regulation of chromatin structure by site-specific histone H3 methyltransferases. Nature. 2000;406(6796):593–9.

32. Zhang M, Liang C, Chen Q, Yan H, Xu J, Zhao H, et al. Histone H2A phosphorylation recruits topoisomerase IIalpha to centromeres to safeguard genomic stability. EMBO J. 2020;39(3):e101863.

33. Kerpel-Fronius S, Gyergyay F, Hindy I, Decker A, Sawinsky I, Faller K, et al. Phase I-II trial of aclacinomycin A given in a four-consecutive-day schedule to patients with solid tumours. A South-East European Oncology Group (SEEOG) Study. Oncology. 1987;44(3):159–63.

34. Martino S, Decker DA, Hynes HE, Kresge CL. Phase II evaluation of aclacinomycin-A in advanced ovarian carcinoma. Investigational new drugs. 1987;5(4):373–4.

35. Johnson SA, Richardson DS. Anthracyclines in haematology: pharmacokinetics and clinical studies. Blood Rev. 1998;12(1):52–71.

36. Fujihira S, Yamamoto T, Matsumoto M, Yoshizawa K, Oishi Y, Fujii T, et al. The high incidence of atrial thrombosis in mice given doxorubicin. Toxicol Pathol. 1993;21(4):362–8.

37. Lencova-Popelova O, Jirkovsky E, Mazurova Y, Lenco J, Adamcova M, Simunek T, et al. Molecular remodeling of left and right ventricular myocardium in chronic anthracycline cardiotoxicity and post-treatment follow up. PloS one. 2014;9(5):e96055.

38. Zhao S, Wu H, Xia W, Chen X, Zhu S, Zhang S, et al. Periostin expression is upregulated and associated with myocardial fibrosis in human failing hearts. J Cardiol. 2014;63(5):373–8.

39. Guo R, Hua Y, Ren J, Bornfeldt KE, Nair S. Cardiomyocyte-specific disruption of Cathepsin K protects against doxorubicin-induced cardiotoxicity. Cell Death Dis. 2018;9(6):692.

40. Lotrionte M, Biondi-Zoccai G, Abbate A, Lanzetta G, D’Ascenzo F, Malavasi V, et al. Review and meta-analysis of incidence and clinical predictors of anthracycline cardiotoxicity. The American journal of cardiology. 2013;112(12):1980–4.

41. Ganzel C, Sun Z, Cripe LD, Fernandez HF, Douer D, Rowe JM, et al. Very poor long-term survival in past and more recent studies for relapsed AML patients: The ECOG-ACRIN experience. Am J Hematol. 2018;93(8):1074–81.

42. Dohner H, Estey E, Grimwade D, Amadori S, Appelbaum FR, Buchner T, et al. Diagnosis and management of AML in adults: 2017 ELN recommendations from an international expert panel. Blood. 2017;129(4):424–47.

43. Yamada K, Furusawa S, Saito K, Waga K, Koike T, Arimura H, et al. Concurrent use of granulocyte colony-stimulating factor with low-dose cytosine arabinoside and aclarubicin for previously treated acute myelogenous leukemia: a pilot study. Leukemia. 1995;9(1):10–4.

44. Wei G, Ni W, Chiao JW, Cai Z, Huang H, Liu D. A meta-analysis of CAG (cytarabine, aclarubicin, G-CSF) regimen for the treatment of 1029 patients with acute myeloid leukemia and myelodysplastic syndrome. Journal of hematology & oncology. 2011;4:46.

45. Jin J, Chen J, Suo S, Qian W, Meng H, Mai W, et al. Low-dose cytarabine, aclarubicin and granulocyte colony-stimulating factor priming regimen versus idarubicin plus cytarabine regimen as induction therapy for older patients with acute myeloid leukemia. Leuk Lymphoma. 2015;56(6):1691–7.

46. Owattanapanich W, Owattanapanich N, Kungwankiattichai S, Ungprasert P, Ruchutrakool T. Efficacy and Toxicity of Idarubicin Versus High-dose Daunorubicin for Induction Chemotherapy in Adult Acute Myeloid Leukemia: A Systematic Review and Meta-analysis. Clin Lymphoma Myeloma Leuk. 2018;18(12):814–21 e3.

47. Wang H, Xiao X, Xiao Q, Lu Y, Wu Y. The efficacy and safety of daunorubicin versus idarubicin combined with cytarabine for induction therapy in acute myeloid leukemia: A meta-analysis of randomized clinical trials. Medicine (Baltimore). 2020;99(24):e20094.

48. McGowan JV, Chung R, Maulik A, Piotrowska I, Walker JM, Yellon DM. Anthracycline Chemotherapy and Cardiotoxicity. Cardiovasc Drugs Ther. 2017;31(1):63–75.

49. DiNardo CD, Tiong IS, Quaglieri A, MacRaild S, Loghavi S, Brown FC, et al. Molecular patterns of response and treatment failure after frontline venetoclax combinations in older patients with AML. Blood. 2020;135(11):791–803.

50. Wu JF, Zhou JJ, Li XA, Hu LH, Wen ML. The safety and efficacy of amrubicin in the treatment of previously untreated extensive-disease small-cell lung cancer: a meta-analysis. Onco Targets Ther. 2019;12:5135–42.

51. DiNardo S, Voelkel K, Sternglanz R. DNA topoisomerase II mutant of Saccharomyces cerevisiae: topoisomerase II is required for segregation of daughter molecules at the termination of DNA replication. Proceedings of the National Academy of Sciences of the United States of America. 1984;81(9):2616–20.

52. Uemura T, Yanagida M. Isolation of type I and II DNA topoisomerase mutants from fission yeast: single and double mutants show different phenotypes in cell growth and chromatin organization. EMBO J. 1984;3(8):1737–44.

53. Uemura T, Ohkura H, Adachi Y, Morino K, Shiozaki K, Yanagida M. DNA topoisomerase II is required for condensation and separation of mitotic chromosomes in S. pombe. Cell. 1987;50(6):917–25.

54. Wander DPA, van der Zanden SY, Vriends MBL, van Veen BC, Vlaming JGC, Bruyning T, et al. Synthetic (N,N-Dimethyl)doxorubicin Glycosyl Diastereomers to Dissect Modes of Action of Anthracycline Anticancer Drugs. The Journal of organic chemistry. 2021;86(8):5757–70.

55. Holm C, Goto T, Wang JC, Botstein D. DNA topoisomerase II is required at the time of mitosis in yeast. Cell. 1985;41(2):553–63.

56. McPherson JP, Goldenberg GJ. Induction of apoptosis by deregulated expression of DNA topoisomerase IIalpha. Cancer research. 1998;58(20):4519–24.

57. Seattle IfHMaEI, United States. Global Burden of Disease Study 2019 Results. Global Burden of Disease Collaborative Network. 2020: Available from https://vizhub.healthdata.org/gbd-results/.

