## Supplementary Materials for "Diverging the anthracycline class of anti-cancer drugs for superior survival of acute myeloid leukemia patients"

Xiaohang Qiao *et.al.*

Jacques Neefjes,

**The Word file includes:**

Figures S1 to S5

Tables S1 to S2

**Supplemental Figures**

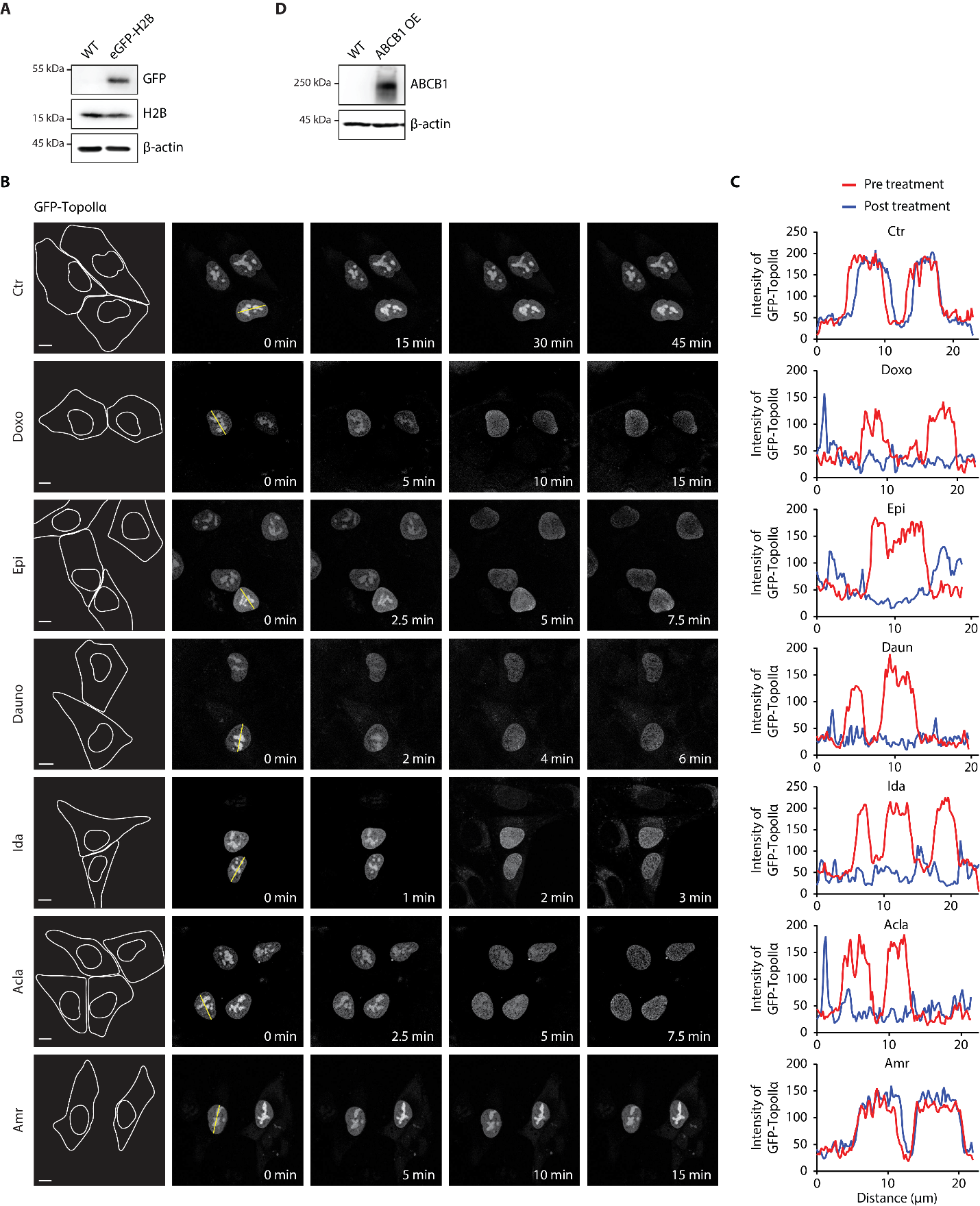

**Figure S1. Histone-evicting anthracyclines redistribute TopoIIα.** (A) Western blot confirmation of K562-eGFP-H2B cells. (B) Redistribution of GFP-TopoIIα imaged by time-lapse confocal microscopy. Snapshots of indicated time points. Left panel, position of cell and nucleus. Scale bar, 10 μm. (C) Localization analysis of GFP-TopoIIα using line intensity profiles. (D) Western blot confirmation of ABCB1-overexpressing K562 cells.

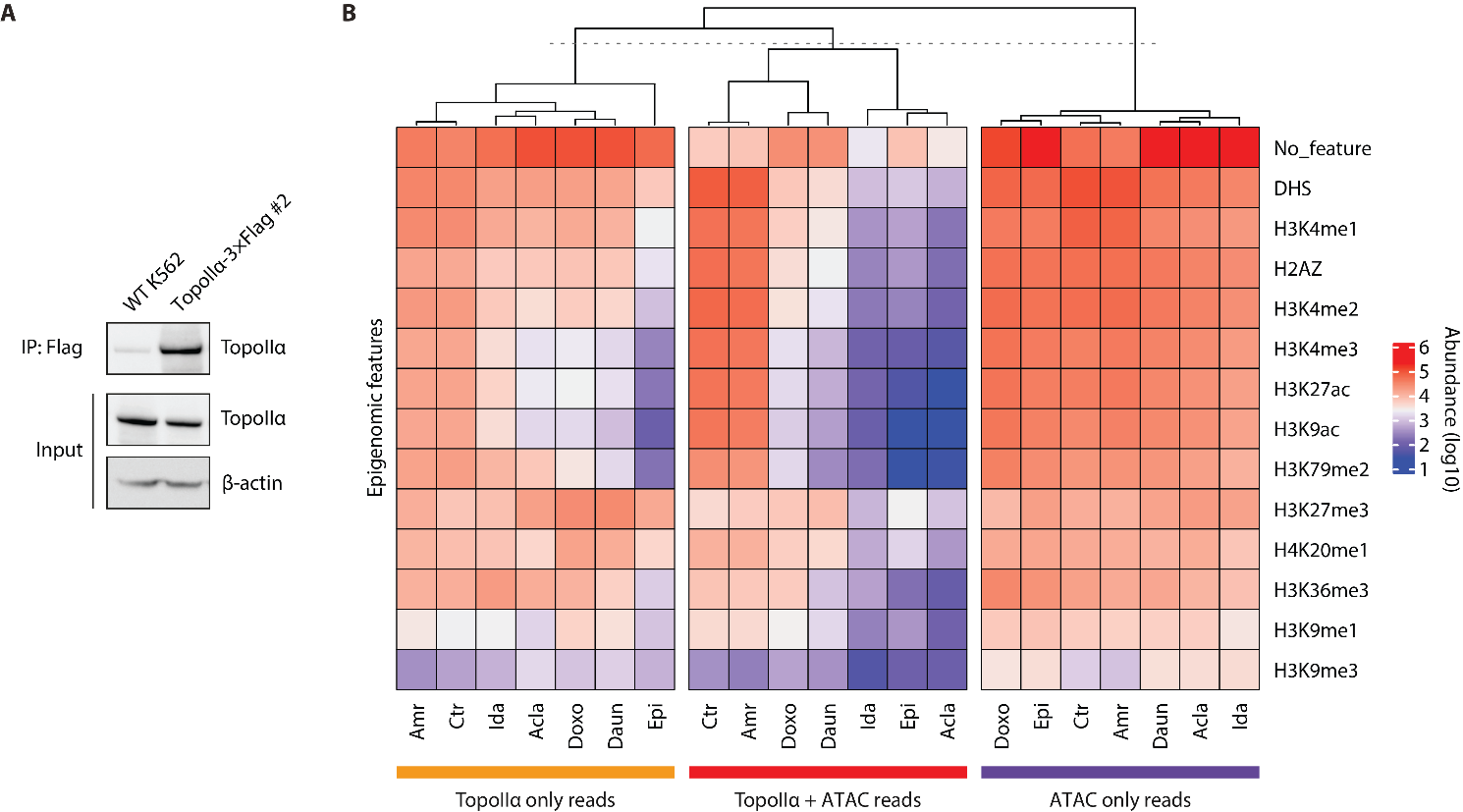

**Figure S2. Epigenetic selectivity of TopoIIα redistribution and histone eviction of anthracyclines.** (A) Western blot confirmation of endogenously tagged TopoIIα-3×Flag K562 cells. (B) Heatmap of peak abundance of each category associated with specific histone features derived from Roadmap Epigenomics Project.

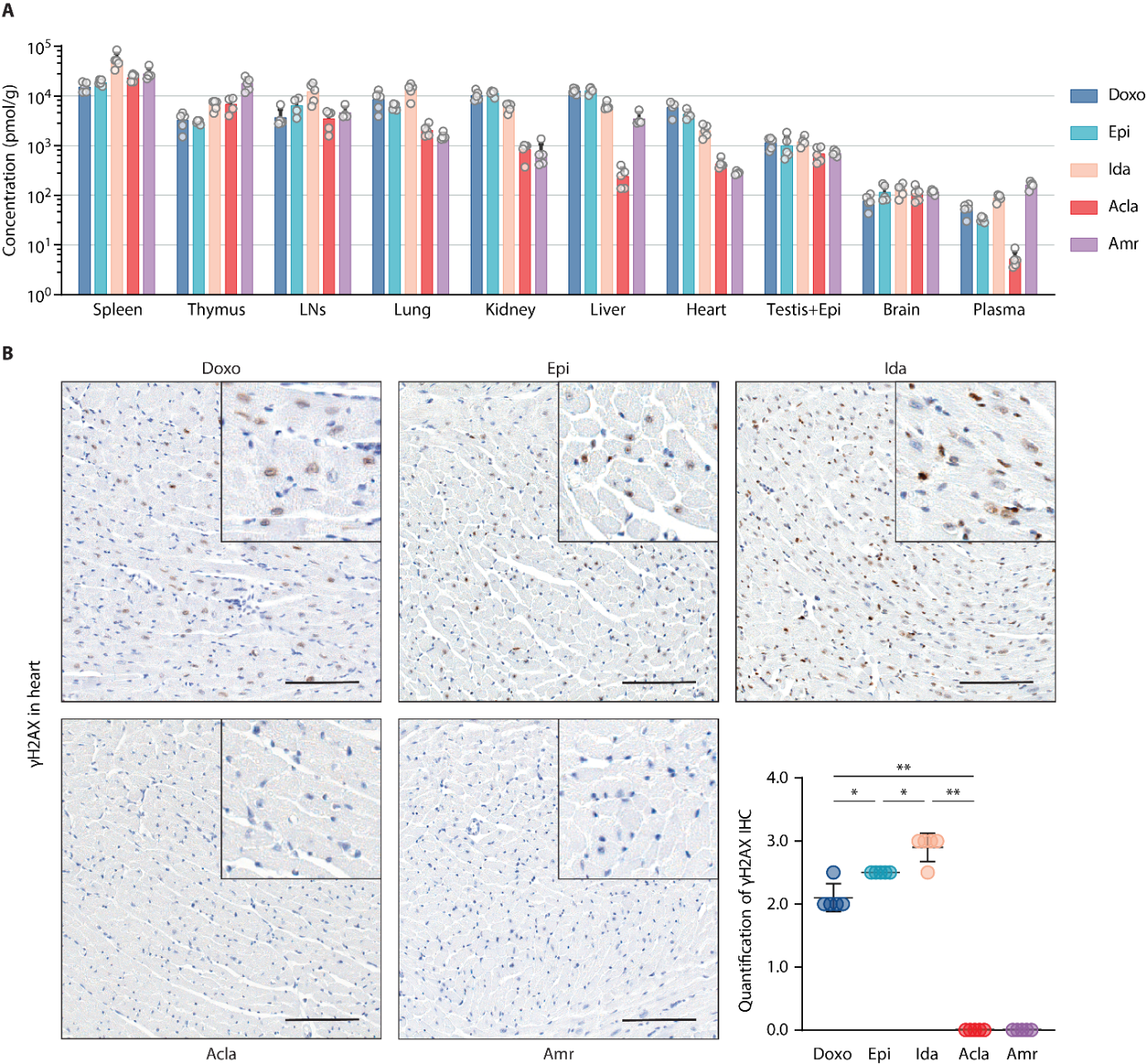

**Figure S3. Bio-distribution and effects of clinically-used anthracyclines in mice.** (A) Drug bio-distribution was determined 4 hours after i.v. injection at 5 mg/kg (*n* = 5 per group). Data are represented as mean ± SD, Student’s *t*-test. (B) Representative microscopic images of γH2AX IHC staining of the hearts from the mice treated in (A). Scale bars, 100 μm. Quantification is represented as mean ± SD, Mann-Whitney test. **P* < 0.05 and ***P* < 0.01.

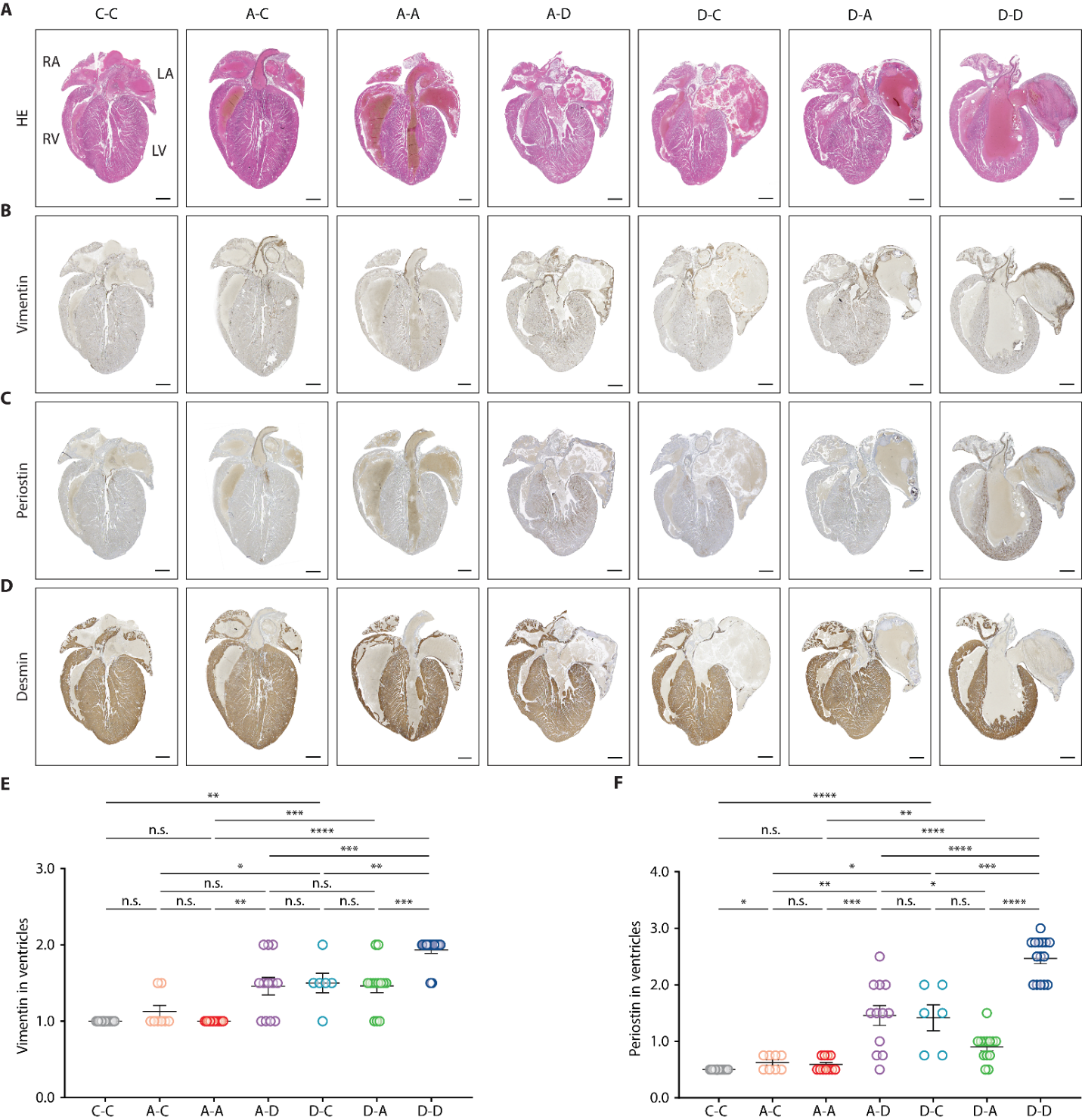

**Figure S4. Acla is safe and well tolerated after Doxo treatment.** FVB mice were treated as in Fig.4. (A‒D) Representative microscopic images of the heart IHC. Scale bars, 500 μm. (E,F) Quantification of the indicated IHC staining in the ventricles of heart. Data are mean ± SEM, Mann-Whitney test.

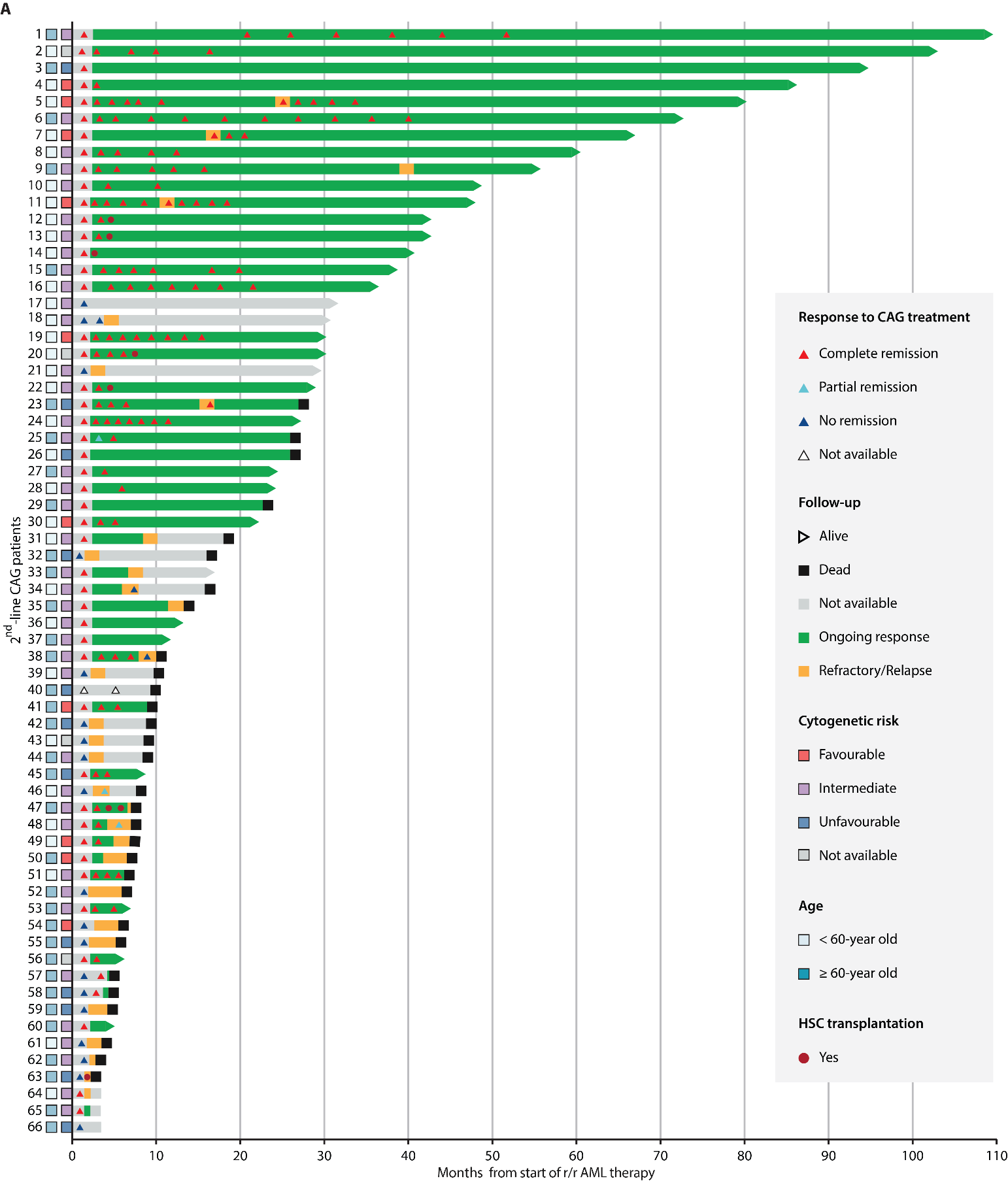

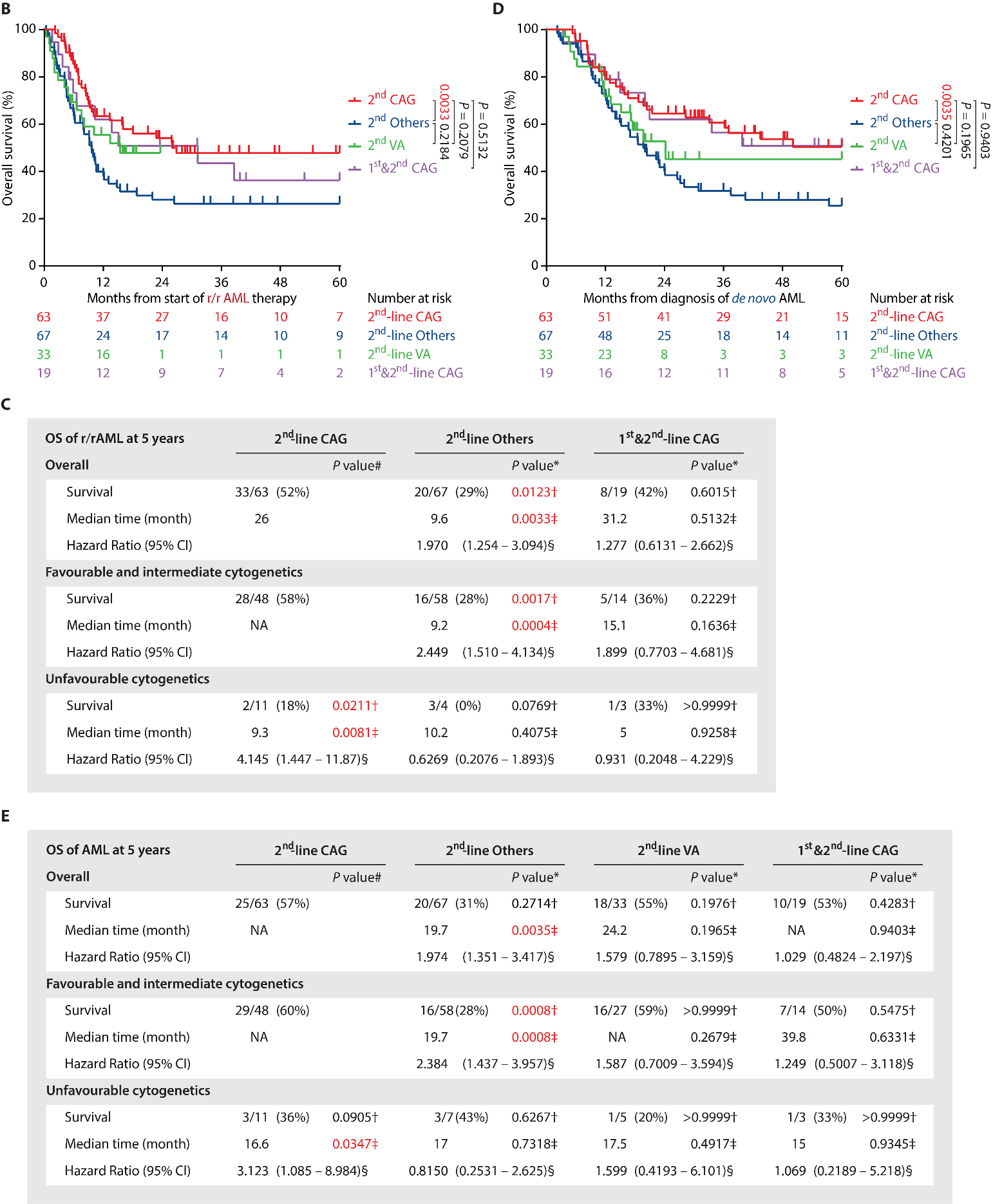

**Figure S5. CAG is effective in treating r/rAML patients.** (A) Swimmer plot showing the remission and survival status of individual 2^nd^-line CAG patients. (B) Overall survival of the r/rAML patients from the start of induction treatment to the date of death. (C) The statistical analysis of OS of r/rAML patients at 5 years (B). Data are n/N (%), * compared to 2^nd^-line CAG group, † Fisher’s exact test, ‡ Log-rank test, § Mantel-Haenszel test, # compared to 2^nd^-line CAG patients with favorable and intermediate cytogenetics. (D) Overall survival of the AML patients from the time of primary disease diagnosis to the date of death. (E) The statistical analysis of OS of AML patients at 5 years (D). Data are n/N (%), * compared to 2^nd^-line CAG group, † Fisher’s exact test, ‡ Log-rank test, § Mantel-Haenszel test, # compared to 2^nd^-line CAG patients with favorable and intermediate cytogenetics.

**Supplemental Tables**

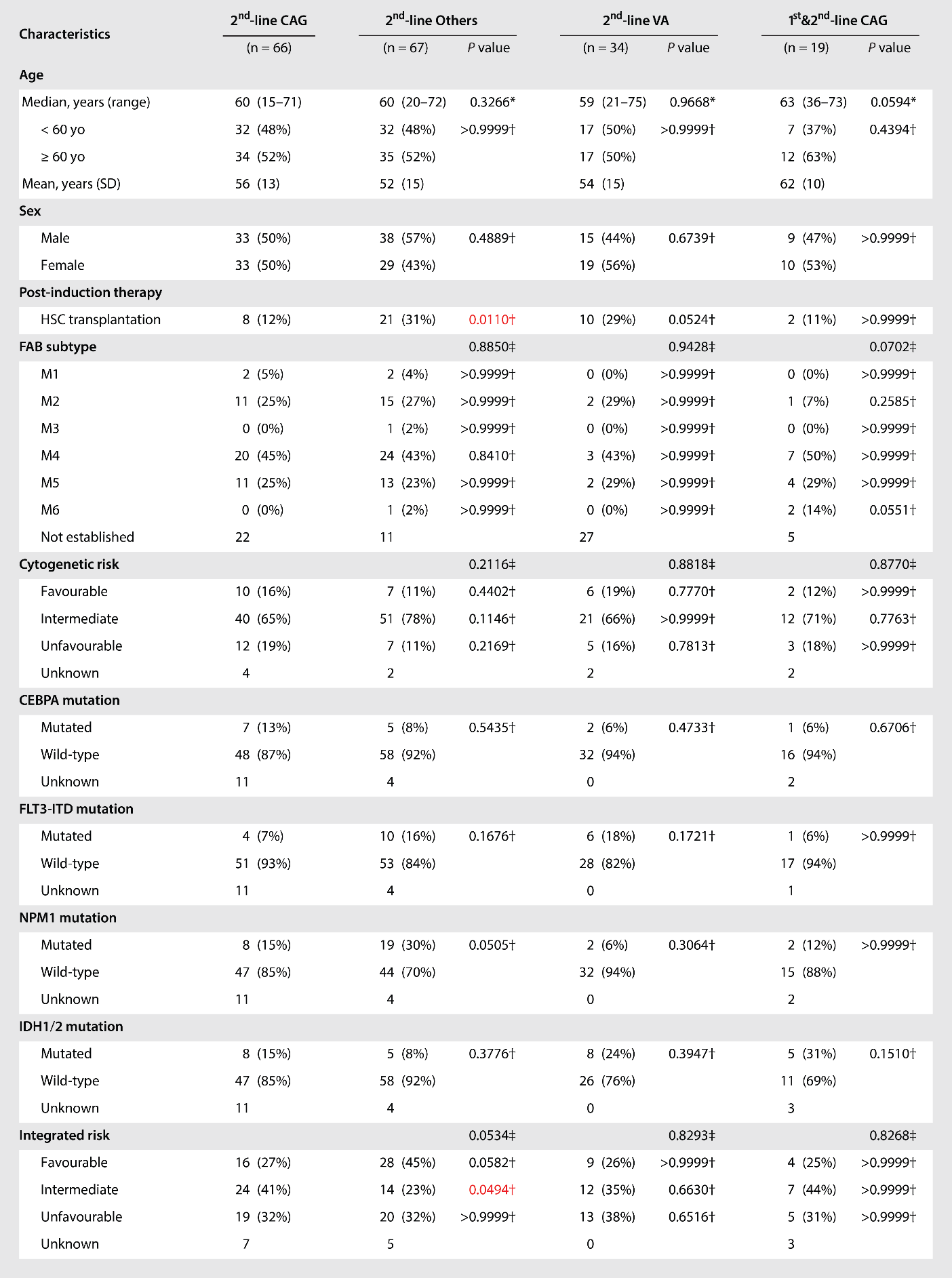

**Table S1. Characteristics of r/rAML patients.** CAG: cytarabin, aclarubicin, G-CSF; IA: idarubicin, cytarabine; VA: venetoclax, azacitidine; Others: chemotherapy regimens other than CAG. Data are n/N (%). † Fisher's exact test between the indicated group and 2^nd^-line CAG group. ‡ Fisher's exact test within group.

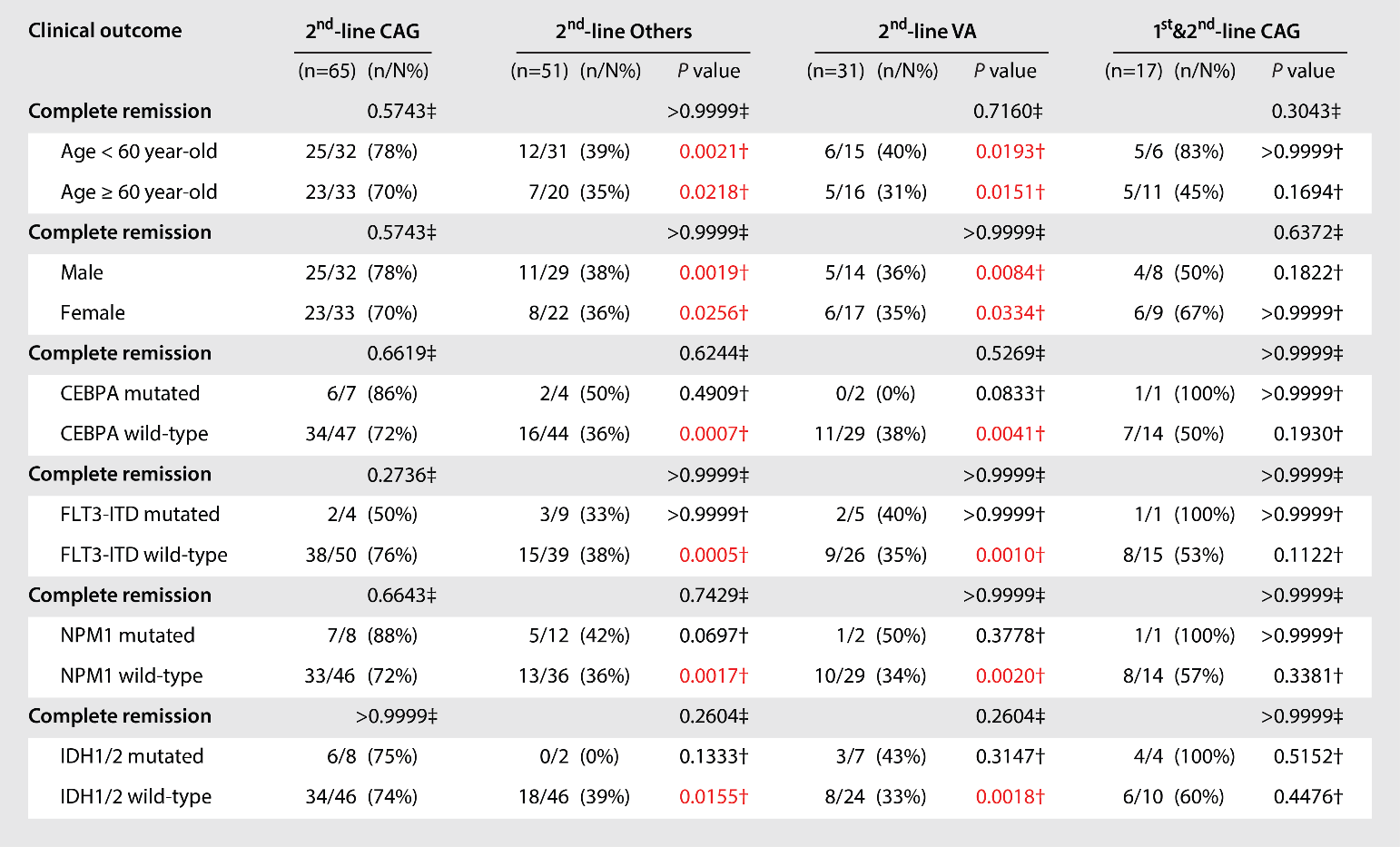

**Table S2. Treatment outcomes of r/rAML patients.** CAG: cytarabin, aclarubicin, G-CSF; IA: idarubicin, cytarabine; VA: venetoclax, azacitidine; Others: chemotherapy regimens other than CAG. Data are n/N (%). † Fisher's exact test between the indicated group and 2^nd^-line CAG group. ‡ Fisher's exact test within group.
